# Common and rare genetic variants show network convergence for a majority of human traits

**DOI:** 10.1101/2025.06.27.25330419

**Authors:** Sarah N. Wright, Jane Yang, Trey Ideker

## Abstract

While both common and rare variants contribute to the genetic etiology of complex traits, whether their impacts manifest through the same effector genes and molecular mechanisms is not well understood. Here, we systematically analyze common and rare variants associated with each of 373 phenotypic traits within a large biological knowledge network of gene and protein interactions. While common and rare variants implicate few shared genes, they converge on shared molecular networks for more than 75% of traits. We demonstrate that the strength of this convergence is influenced by core factors such as trait heritability, gene mutational constraints, and tissue specificity. Using neuropsychiatric traits as examples, we show that common and rare variants impact shared functions across multiple levels of biological organization. These findings underscore the importance of integrating variants across the frequency spectrum and establish a foundation for network-based investigations of the genetics of diverse human diseases and phenotypes.

## INTRODUCTION

Genome-wide association studies (GWAS) have identified thousands of common variants (CVs) that are significantly associated with complex human traits, including a wide array of diseases and clinical biomarkers^1^. However, elucidating causal mechanisms impacted by these CVs has proven challenging because they typically fall in non-coding regions and exert small effect sizes. Therefore, GWAS have been recently complemented by large-scale exome and whole-genome sequencing efforts, which have enabled the identification of trait-associated variants present at low frequencies in coding regions^2^. The resulting rare variants (RVs) exert larger effect sizes^3^ and directly implicate genes and biological pathways^4^. Separately, these common and rare variant association studies have identified important genetic factors underlying human diseases and other complex traits^1^; however, their joint influence remains poorly understood.

There is growing evidence that RVs contribute to trait heritability and that incorporating them alongside CVs can improve polygenic risk score (PRS) predictions^3,5^. However, the contribution of RVs appears to be highly trait-dependent. For example, Wainschtein et al. found that RVs in low linkage disequilibrium regions significantly drive variance in human height, and Fiziev et al. showed that rare-variant PRS are better at identifying individuals at phenotypic extremes across 78 phenotypes. Conversely, minimal contributions from RVs have been reported for the polygenic risk prediction of metabolic traits^6^. Moreover, global heritability and polygenic risk score analyses do not necessarily provide insights in to the biological mechanisms impacted by genetic variation.

Previous studies have shown that CV and RV associations can systematically prioritize different genetic loci and effector genes^7^. These differences have been linked to challenges such as limited statistical power, differing selection pressures, population stratification, or heterogeneity in clinical manifestations^8–10^. CVs and RVs at different genetic loci, however, do not necessarily imply functional divergence within a trait. For example, for schizophrenia^11^ and autism spectrum disorder^12^ such variants have been shown to converge onto relevant molecular networks that implicate protein complexes and biological pathways. However, to date, such systems-level concordance has been demonstrated in only a small number of human phenotypes.

Here, we examine the role of common and rare variants across the human phenome via a systematic network integration of CVs and RVs for 373 distinct traits. We identify significant network convergence for 283 phenotypes, 84 of which have no shared common and rare variant-associated genes. In so doing, we aim to understand the factors that drive differences in the discovered CVs and RVs, as well as their overall network convergence.

## RESULTS

### Identification of 373 human traits with common and rare variants results

We identified phenotypes and common and rare variant associations from studies curated by the GWAS Catalog^13^ and the Rare Variant Association Repository^14^ (RAVAR) (**Figure 1A**). Common-variant associated genes (CVGs) were defined as distinct genes with a trait-associated single-nucleotide polymorphism (SNP) located within the gene region defined by Ensembl^13,15^. After quality control (Methods), we identified 7538 studies with at least three suggestive CVGs (*p* < 1×10^−5^), representing 2339 distinct traits. Separately, we sourced rare variant-associated genes (RVGs) from gene-based rare variant studies in the RAVAR database. We identified 2207 studies representing 1121 distinct traits with at least three suggestive RVGs (*p* < 1×10^−4^). From these studies, we identified 373 traits with both common and rare variant association results.

**Figure 1.**
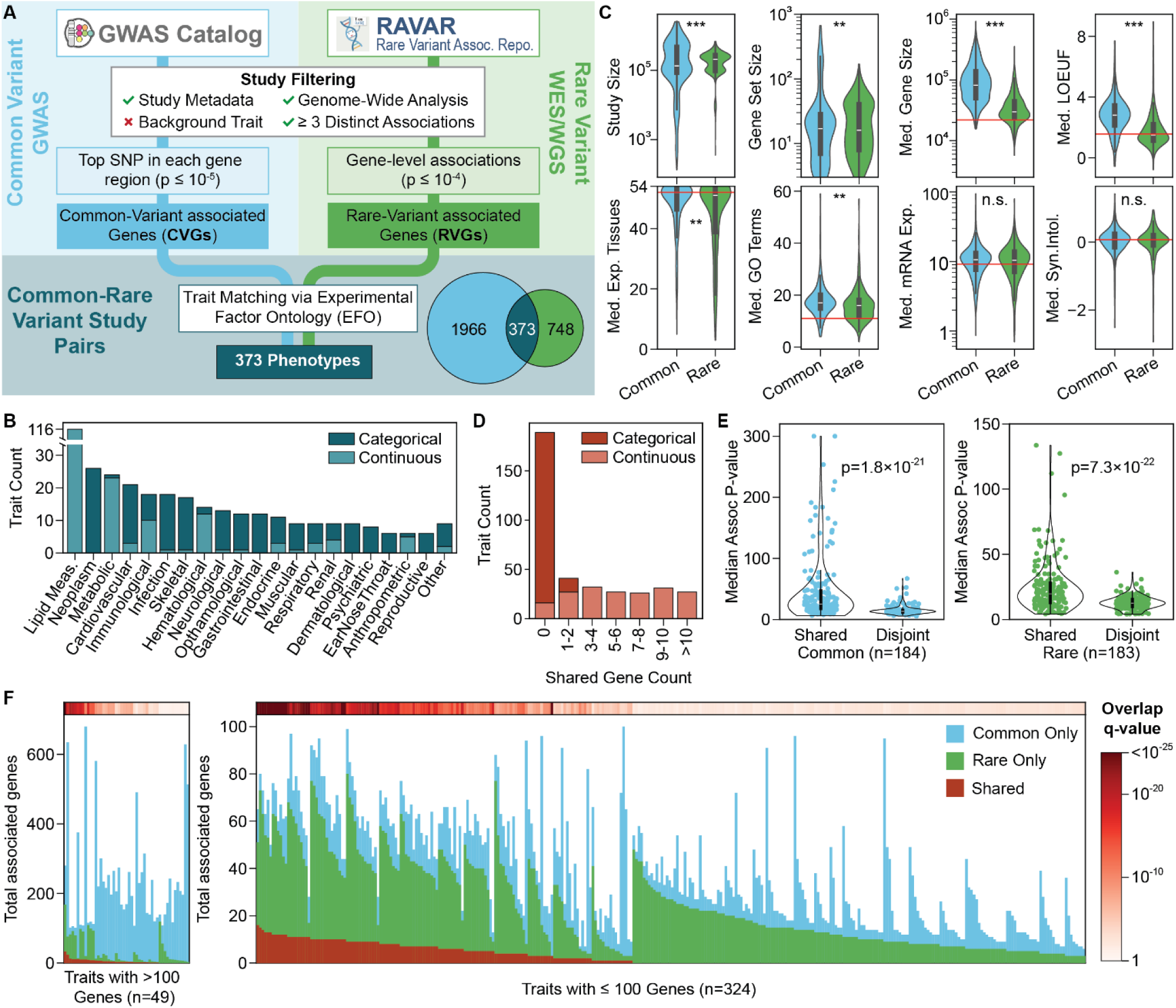
Curation and analysis of common and rare variant associations for 373 traits. **A)** Overview of the data curation and trait matching pipeline for defining sets of common-variant associated genes (CVGs) and rare-variant associated genes (RVGs). **B)** Distribution of traits across biological domains, stacked by trait classifications. Domains with fewer than five traits are included in ‘Other’. **C)** Comparison of CVG and RVG properties across 373 traits (Wilcoxon Signed-Rank Test, BH correction). For gene-level properties, the value for each set of CVGs or RVGs was taken as the median of all set genes. The center bar represents the median, the box represents the interquartile range (Q1–Q3), and the upper and lower whiskers represent Q1 − 1.5IQR and Q3 + 1.5IQR. The violins extend to the minimum and maximum observations. Red lines indicate the median value for all protein-coding genes, where applicable. *** *q* < 1×10^−5^, ** *q* < 1×10^−3^, n.s. *q* > 0.05. **D)** Distribution of the number of shared genes identified per trait as both CVGs and RVGs, stacked by trait classification. **E)** Comparison of the median CVG (left) and RVG (right) association p-values between shared genes and disjoint genes (Wilcoxon Signed-Rank test). The center bar represents the median, the box represents the interquartile range (Q1–Q3), and the upper and lower whiskers represent Q1 − 1.5IQR and Q3 + 1.5IQR. The violins extend to the minimum and maximum observations. **F)** Stacked bar chart of the total number of distinct genes associated with each trait. Shared genes are those identified as both CVGs and RVGs. The top heatmap shows the q-value of the number of shared genes (hypergeometric test, BH correction). Plot split into traits with > 100 associated genes and traits with ≤ 100 associated genes for visualization. See also Supplemental Tables 1 & 2.

This collection of traits represented a variety of clinical biomarkers, disease states, and other complex phenotypes (**Figure 1B**). Among categorical traits (n = 187), neoplasms and cardiovascular diseases were the most represented domains, while lipid and other metabolic measurements were the most represented domains among continuous traits (n = 186). Study populations varied from large biobank cohorts, such as the UK Biobank, to smaller ascertained studies. Rare variant study populations tended to be larger, with a median of 210,000 individuals, compared to a median of 136,000 individuals for common variant studies (**Figure 1C**, *q* = 6.8×10^−8^). We also observed that RVGs were more mutationally constrained (*q* = 6.2×10^−33^), as evidenced by lower median loss-of-function observed to expected upper bound fractions (LOEUF) – a continuous metric that measures a gene’s intolerance to loss-of-function variation^16^. In contrast, CVGs had longer gene regions (*q* = 1.5×10^−50^) and more functional annotations (*q* = 2.7×10^−5^), with expression in a greater number of tissues (*q* = 5.5×10^−4^).

### Common and rare variants implicate few shared genes

Identifying a gene via both CV and RV analyses can lend confidence to gene-trait associations. Overall, continuous traits identified a greater number of CV/RV shared genes than categorical traits (**Figure 1D**). The greatest agreement was observed for HDL cholesterol change measurement, with 11 shared genes among 22 CVGs and 17 RVGs (*q* = 5.4×10^−32^). Comparing the association p-values of shared genes to all common-only and rare-only associations, we found that the shared gene associations tended to be more significant than the disjoint associations (**Figure 1E**), supporting their role as high-confidence trait associations. While 184 traits had significantly more shared genes than expected by chance (**Figure 1F**), these shared genes represented a minority of all CVGs and RVGs (median of 28% and 18%, respectively). Furthermore, of the 373 phenotypes, 189 did not identify any shared genes. The minimal correspondence between CVGs and RVGs for a large proportion of traits suggested that focusing solely on shared genes does not provide a complete picture of trait genetics.

### A majority of traits demonstrate network convergence of common and rare variants

To move beyond individual genes and investigate shared functions, we integrated CVGs and RVGs within a comprehensive biological knowledge network, the Parsimonious Composite Network (PCNet2), which contains 3.85 million pairwise relationships among human genes and proteins^17^. Using an approach called Network Colocalization^18,19^ (NetColoc, **Figure 2A**), we propagated the CVG and RVG association scores throughout the network. From each trait’s propagated scores, we formed trait networks that captured the subset of genes in close network proximity to both CVGs and RVGs. To quantify overall network convergence, we defined the COLOC score as the observed over expected size of the trait network.

**Figure 2.**
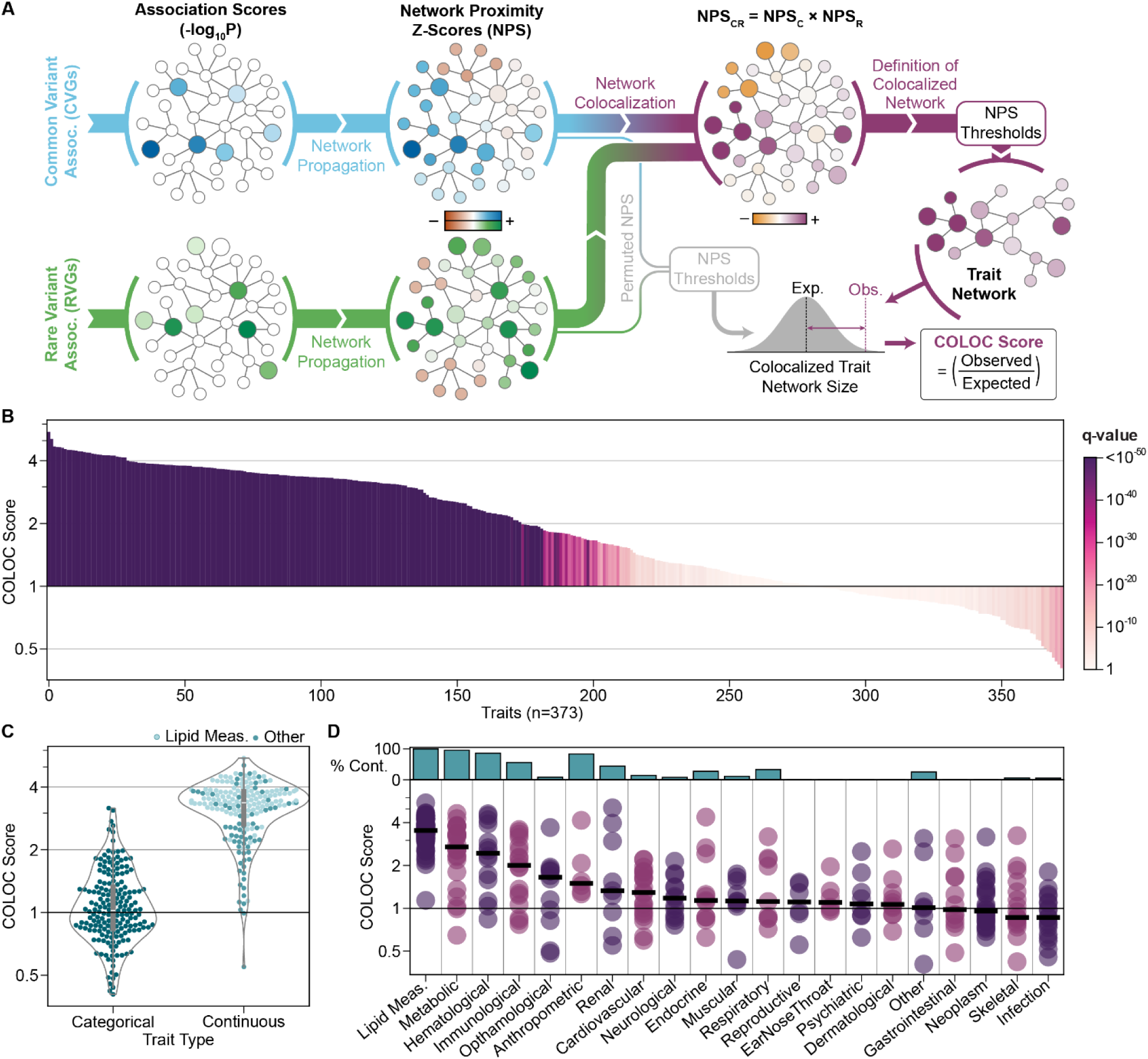
Network convergence of common and rare variants across 373 human traits. **A)** Schematic of the Network Colocalization (NetColoc) pipeline for assessing the convergence of CV and RV associated genes and defining colocalized trait networks. **B)** Network colocalization of common and rare variants for 373 traits. Traits are ordered by COLOC score, and bar color indicates q-value of the COLOC score based on 1000 permutations with BH-correction. **C)** Comparison of COLOC scores between categorical and continuous traits. The center bar represents the median, the box represents the interquartile range (Q1−Q3), and the upper and lower whiskers represent Q1 − 1.5IQR and Q3 + 1.5IQR. **D)** Distribution of COLOC scores across biological domains. Black lines show the median COLOC score per domain. The upper bar plot shows the percentage of domain traits that are continuous. See also Supplemental Table 2.

Of the 373 traits, 254 showed significant network convergence of CVGs and RVGs (**Figure 2B**), suggesting a high concordance in the molecular systems impacted by common and rare variants. Of these, 174 traits produced trait networks at least 2 times larger than expected by chance (COLOC score ≥ 2). Continuous traits showed significantly stronger CVG/RVG network convergence, consistent with the higher number of shared genes observed for these traits (**Figure 2C**, *p* = 1.0×10^−54^). Network convergence was particularly strong among lipid measurement traits, which had trait networks that were, on average, 3.6 times larger than expected by chance. In contrast, infection, neoplasm, and skeletal traits exhibited low rates of network convergence, with significant COLOC scores for only 4 of 18, 9 of 26, and 6 of 17 traits, respectively (**Figure 2D**).

### Network convergence is robust across variable study designs

To test generalizability across studies, we performed additional NetColoc analyses using 223 of the 373 traits for which multiple studies of common or rare variants were available. Up to four additional CV studies and four additional RV studies were selected per trait (**Figure 3A**), prioritizing those conducted in different population cohorts. COLOC scores for each trait were largely consistent across the various studies (**Figure 3B**, r_s_ = 0.84). Nonetheless, the variation amongst scores highlighted that the strength of CVG/RVG convergence observed can be influenced by study selection. Some traits, such as Crohn’s disease and Cystatin C measurement, showed large discrepancies between different CV studies, while others, such as Schizophrenia and Eosinophil count, showed discrepancies between different RV studies (**Figure 3C**, **Supplemental Figure 1A**).

**Figure 3.**
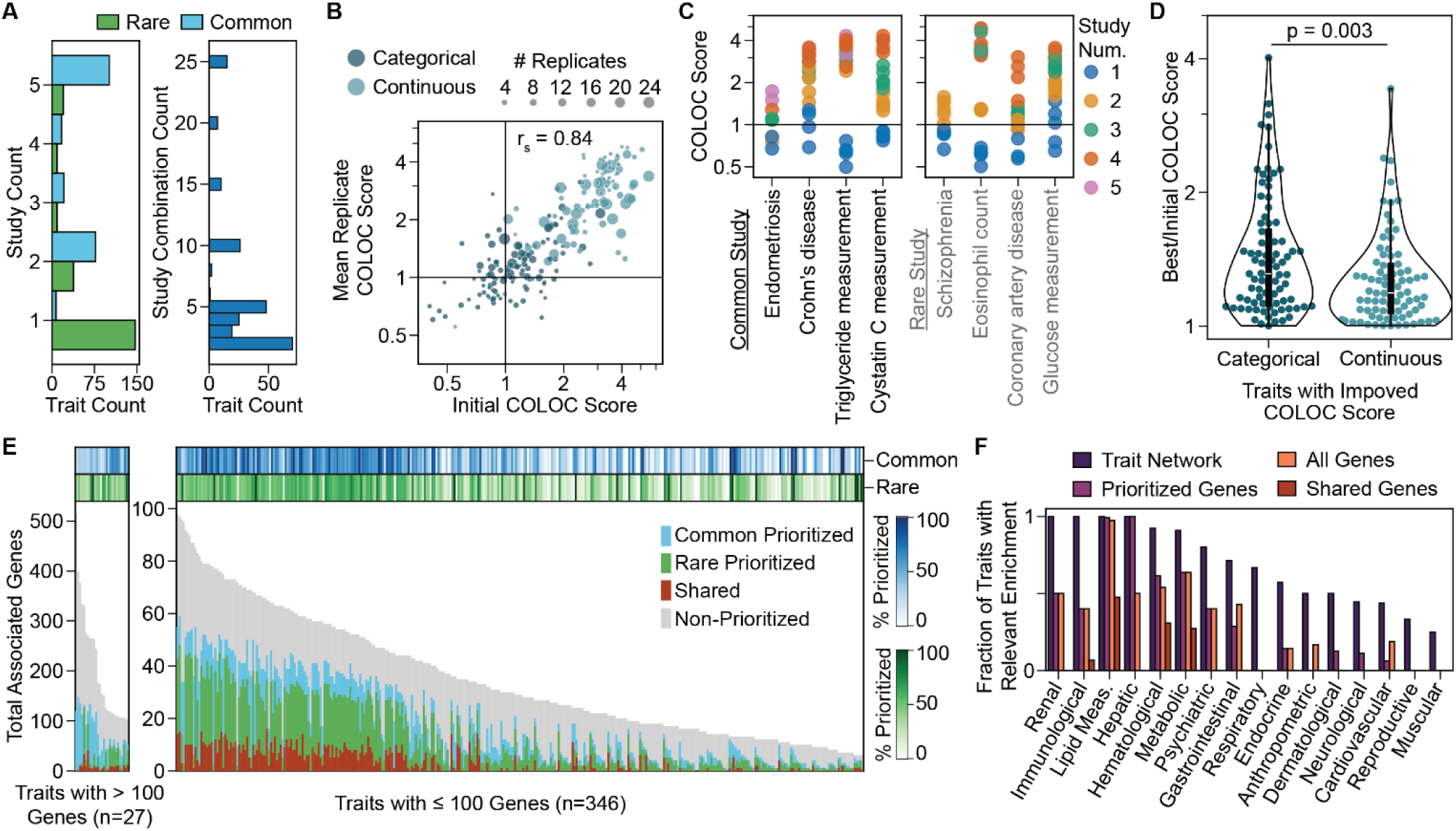
Robustness of network convergence results and prioritization of trait associations. **A)** Total number of additional CV studies and RV studies utilized for robustness testing per trait, up to a maximum of five (left), and total number of common-rare study combinations analyzed per trait (right). Results shown for the 223 traits with at least one additional study. **B)** Initial COLOC scores (Figure 2C), compared to the mean COLOC of all additional analyses for that trait. Color indicates trait classification, and size indicates the number of additional study combinations tested. Results shown for the 223 traits with at least one additional study. **C)** COLOC scores for eight example traits with differential dependence on CV study selection (left) or RV study selection (right). COLOC scores for CV study-dependent traits are colored by the common variant study, and COLOC scores for RV study-dependent traits are colored by the rare variant study. **D)** Ratio of best to initial COLOC scores following identification of the optimal study combination per trait, compared across categorical (n=85) and continuous traits (n=83) via Mann-Whitney U Test. Box plots show the median and interquartile range. Traits with an improved COLOC score are shown. **E)** Stacked bar plot showing the prioritization of gene associations following network colocalization. Prioritized genes were defined as disjoint CVGs or RVGs included in the best-scoring trait network for each trait. Top heat maps show the fraction of all CVGs and RVGs prioritized by the network analysis. **F)** Fraction of traits per domain with at least one significant enrichment for genes expressed in a relevant tissue. Tissue-specific enrichment was performed using Human Protein Atlas (HPA) mRNA expression values for all genes in the trait network (dark purple), prioritized input genes (purple), union of all CVGs and RVGs (orange), or only the shared CVGs and RVGs (red). Relevant tissues were defined manually based on biological domains (Methods). See also Supplemental Figure 1 and Supplemental Table 2.

We assigned the final COLOC score as the maximum score observed for each trait. Despite having fewer total studies, the improvement in score was higher for categorical traits (*p* = 0.003, **Figure 3D**). From these results, we identified significant network convergence for an additional 29 traits (**Supplemental Figure 1B**), bringing the total to 283 traits with high concordance between common and rare variants. Of these, 84 had no shared genes, reinforcing the power of a network approach for integrating non-overlapping, but complementary, gene associations. Overall, 44% of disjoint CVGs and 42% of disjoint RVGs were prioritized based on their global network proximity to other trait-associated genes (**Figure 3E**). Compared to the original input genes, these prioritized trait genes and the full trait networks showed better enrichment for genes expressed in trait-relevant tissues, demonstrating that the network approach prioritizes functionally relevant genes (**Figure 3F**). The network convergence results remained consistent when the network colocalization analysis was performed with three alternative networks, with an average Spearman correlation of 0.92 (**Supplemental Figure 1C**).

### Heritability and gene properties drive network convergence

Heritability and population prevalence are known to impact the discovery of common and rare variants. To examine their impact on the CVG/RVG network convergence, we collected SNP heritability estimates for 197 traits from the UK Biobank^20^, and global prevalence estimates for 87 traits from the Global Burden of Disease study^21^. The COLOC scores for these subsets of traits were strongly correlated with trait heritability (**Figure 4A**, r_s_ = 0.61), but not population prevalence (**Figure 4B**, r_s_ = −0.01).

**Figure 4.**
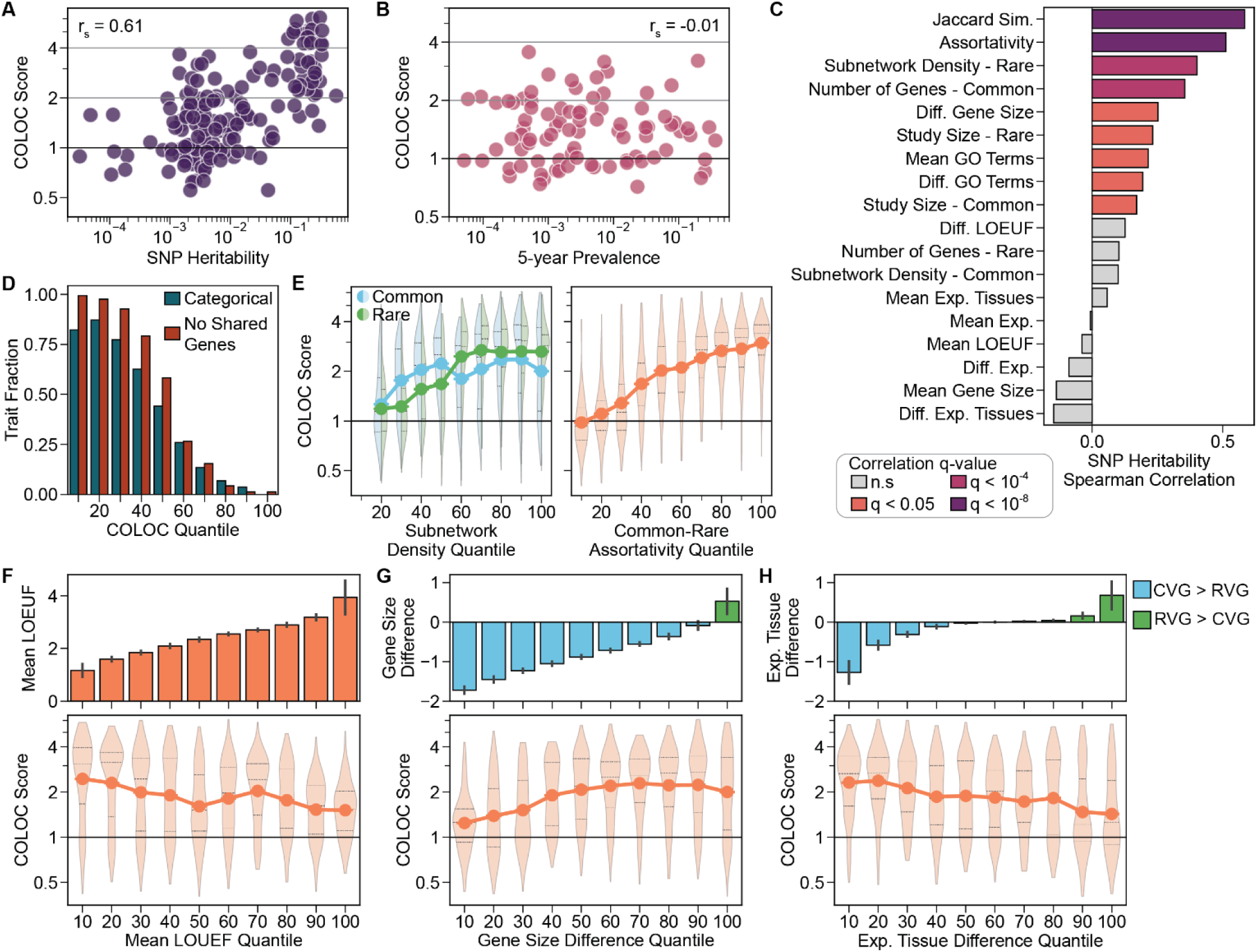
Influence of trait and gene features on the network convergence of common and rare variants. **A-B)** COLOC score as a function of (A) trait SNP heritability (n=197) and (B) trait prevalence (n=87). Spearman correlation reported. Trait prevalence estimates represent averages over the five-year period from 2017 to 2021. **C)** Spearman correlation of trait SNP heritability with 18 trait and gene features. Gene features were summarized by the overall mean of CVGs and RVGs per trait, and the difference between CVGs and RVGs. See also Supplemental Figure 2B. **D)** Fraction of traits within each COLOC score quantile that were categorical traits or had zero shared CVGs and RVGs. **E)** COLOC score as a function of network features of CVGs and RVGs. Point plots show the mean COLOC score for each feature quantile, and violin plots show the distribution of COLOC scores within each feature quantile, with the median and interquartile ranges represented by horizontal lines. Violins extend to the minimum and maximum observed values. **F-H)** COLOC score as a function of (F) Mean LOEUF, (G) Gene size difference, and (H) Expressed tissue difference for CVGs and RVGs. Point plots show the mean COLOC score for each feature quantile, and violin plots show the distribution with median and quartiles as horizontal lines. Upper bar plots show the mean feature value within each feature quantile. Error bars indicate the standard error. For G-H, CVG > RVG indicates that the CVGs had larger gene sizes and were expressed in more tissues, respectively. For D-H, values were calculated for all studies of the 373 traits, including all primary and additional studies to give n = 1629. LOEUF: Loss-of-function intolerance observed/expected upper fraction (mutational constraint). Exp.: median mRNA expression across all tissues. Exp. Tissues: Number of tissues with mRNA expression > 1 TPM. See also Supplemental Figure 2 and Supplemental Tables 3 & 4.

Next, we investigated the influence of study design and gene properties on the extent of network convergence. For all traits, we collected 10 summary features covering the study type and population size, size and network properties of the CVG and RVG sets, and 10 summary values of biological properties, including gene expression, mutational constraints, and functional annotations (**Supplemental Figure 2A-B**). The trait heritability showed widespread correlations with these features (**Figure 4C**), offering potential explanations for how trait heritability mediates the convergence of CVGs and RVGs. For example, traits with higher heritability had a larger number of CVGs and more interconnected RVGs. Using a linear regression model, we found that the trait and gene features explained 77% of the observed variance in COLOC score across all traits, driven by nine significant features (**Table 1**).

**Table 1.**
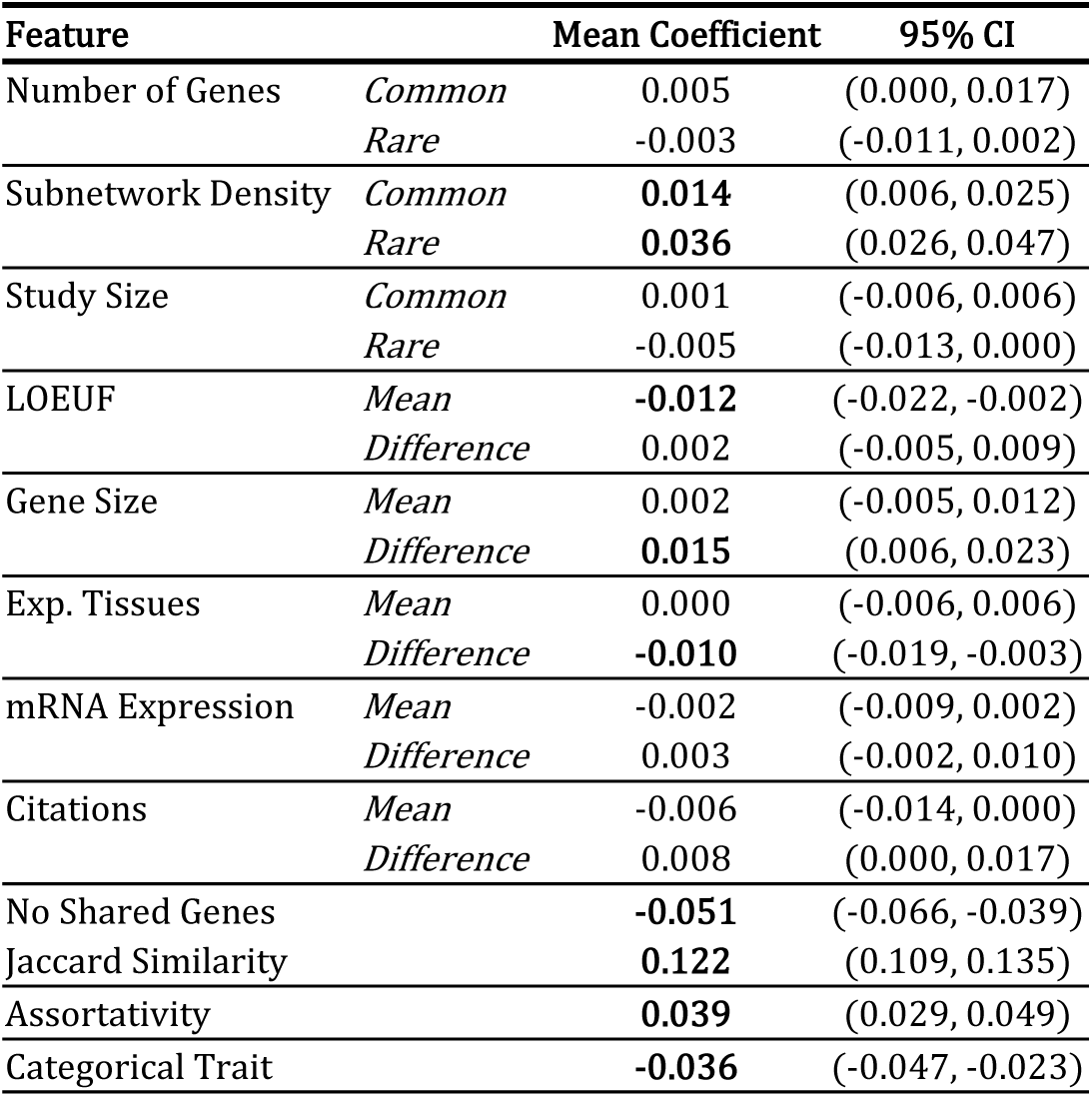
Linear regression estimates of the influence of trait and gene features on COLOC scores. The Elastic Net regression model for predicting COLOC score was optimized via five-fold cross-validation, followed by bootstrap estimation of coefficients using 2000 bootstrap replicates over 1629 samples. Mean coefficient value, 95% confidence interval, and estimated q-value (Bonferroni corrected empirical p-value) are reported. Significant features (*q* < 0.05) are highlighted. LOEUF: Loss-of-function intolerance observed/expected upper fraction (mutational constraint). Exp.: median mRNA expression across all tissues. Exp. Tissues: Number of tissues with mRNA expression > 1 TPM.

| Feature |  | Mean Coefficient | 95% CI |
| --- | --- | --- | --- |
| Number of Genes | *Common* | 0.005 | (0.000, 0.017) |
|  | *Rare* | -0.003 | (-0.011, 0.002) |
| Subnetwork Density | *Common* | **0.014** | (0.006, 0.025) |
|  | *Rare* | **0.036** | (0.026, 0.047) |
| Study Size | *Common* | 0.001 | (-0.006, 0.006) |
|  | *Rare* | -0.005 | (-0.013, 0.000) |
| LOEUF | *Mean* | **-0.012** | (-0.022, -0.002) |
|  | *Difference* | 0.002 | (-0.005, 0.009) |
| Gene Size | *Mean* | 0.002 | (-0.005, 0.012) |
|  | *Difference* | **0.015** | (0.006, 0.023) |
| Exp. Tissues | *Mean* | 0.000 | (-0.006, 0.006) |
|  | *Difference* | **-0.010** | (-0.019, -0.003) |
| mRNA Expression | *Mean* | -0.002 | (-0.009, 0.002) |
|  | *Difference* | 0.003 | (-0.002, 0.010) |
| Citations | *Mean* | -0.006 | (-0.014, 0.000) |
|  | *Difference* | 0.008 | (0.000, 0.017) |
| No Shared Genes |  | **-0.051** | (-0.066, -0.039) |
| Jaccard Similarity |  | **0.122** | (0.109, 0.135) |
| Assortativity |  | **0.039** | (0.029, 0.049) |
| Categorical Trait |  | **-0.036** | (-0.047, -0.023) |

After adjusting for these features, the correlation between trait heritability and COLOC score was reduced (**Supplemental Figure 2C**, r_s_ = 0.16), confirming that the study and gene properties can partly explain the heritability effect. As expected, the number of shared genes, the trait type, and the interconnectedness of the CVGs and RVGs within the network were significant predictors of the network convergence results (**Figure 4D-E**, **Table 1**). Independent of these factors, enhanced convergence was driven by sets of CVGs and RVGs containing more mutationally constrained genes and traits with smaller differences in gene size between CVGs and RVGs (**Figure 4F-G**, **Table 1**). Interestingly, we found that the distribution of gene expression across tissues significantly influenced the observed COLOC score, as measured by the number of tissues with mRNA expression greater than 1 TPM. Traits with RVGs expressed in fewer tissues compared to CVGs showed significantly stronger network convergence (**Figure 4H**, **Table 1**). Other trait and gene set features, such as the number of gene associations, the study population size, and the number of functional annotations, were not independently identified as significant predictors of CVG/RVG network convergence.

### Tissue-specific RVGs are associated with stronger convergence of common and rare variants

Our results indicated that the network convergence of CVGs and RVGs was stronger for traits with RVGs expressed in a limited number of tissues compared to CVGs (**Figure 4H**). To investigate the relevance of these genes, we performed a tissue-specific enrichment analysis for trait-relevant tissues, focusing on the traits exhibiting the largest differences in tissue distribution between the two sets of genes (**Figure 5A-B**). Overall, traits with a significant enrichment for genes expressed in a relevant tissue had higher COLOC scores than traits without such an enrichment (*p* = 0.004, **Figure 5B**). Furthermore, the fraction of gene sets with a relevant enrichment was greater for RVGs, compared to CVGs, indicating that RVGs with limited expression tended to be more consistent with the associated trait.

**Figure 5.**
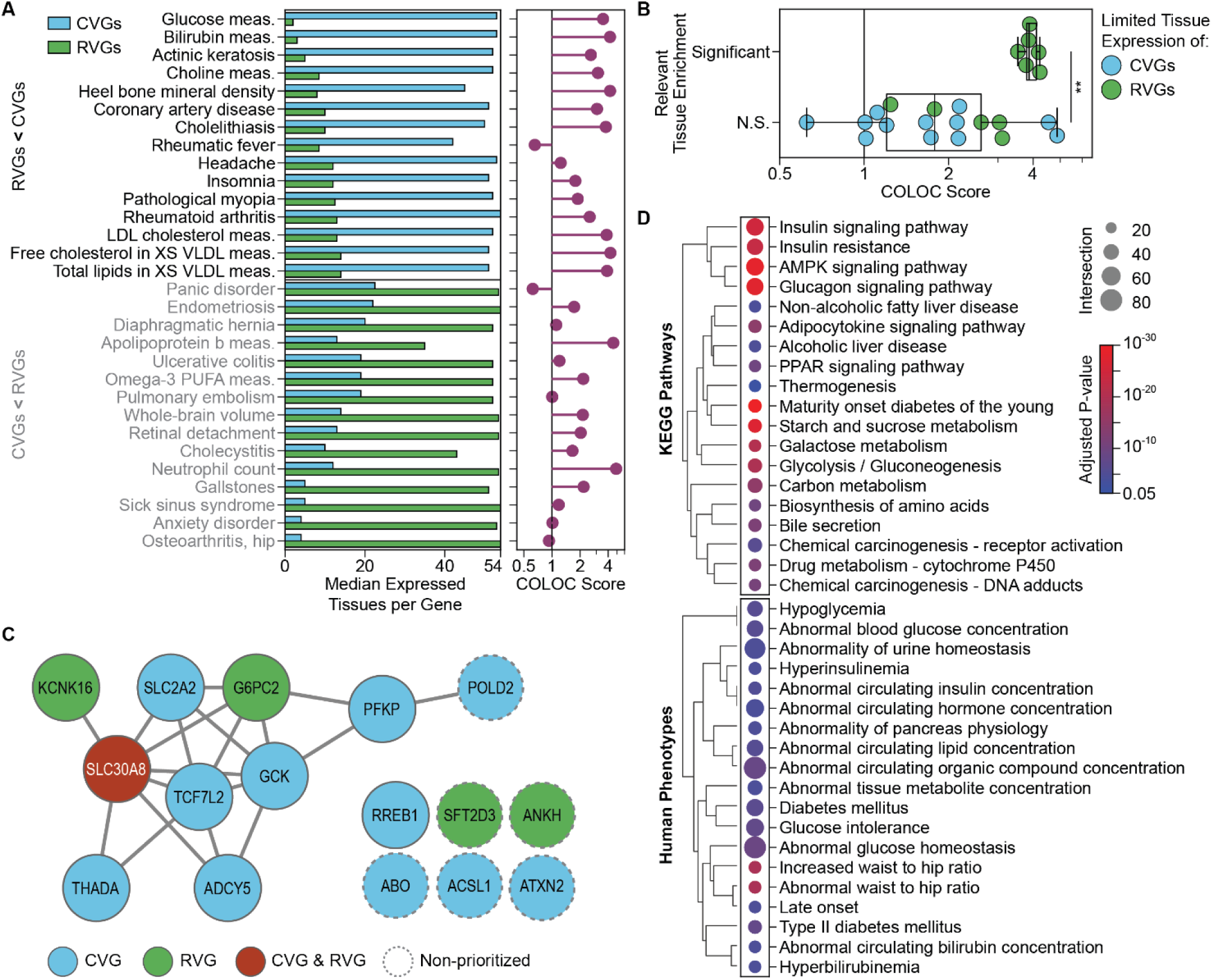
Impacts of gene tissue distribution on the network convergence of common and rare variants. **A)** Tissue distribution of gene expression for CVGs and RVGs, compared to the COLOC score for 30 traits. Bar plot (left) shows the median number of tissues with mRNA expression > 1 TPM for CVGs and RVGs, and lollipop plot (right) shows the corresponding COLOC score for each trait. Traits were selected by the difference between the number of expressed tissues for RVGs and CVGs, including the top 15 traits with CVG > RVG and the top 15 traits with CVG < RVG. **B)** Comparison of the COLOC score to the tissue-specific enrichment in the most relevant tissue (Mann-Whitney U Test, \*\**p* = 0.004). Significant tissue-specific enrichment was defined as an adjusted p-value < 0.05 (Methods) for the gene set (CVG or RVG) with a lower median number of expressed tissues. Results shown for 24 of the top 30 traits in (A) for which a trait-relevant tissue could be defined. **C)** Subnetwork of initial CVGs and RVGs for glucose measurement. Edges derived from PCNet2.0. Non-prioritized genes are those not included in the glucose measurement trait network. **D)** KEGG pathway (top) and Human Phenotype Ontology (HPO, bottom) enrichments for the glucose measurement trait network. Enrichments were filtered to those with adjusted p-value < 0.05 (KEGG) or 5×10^−5^ (HPO), term size < 1000, and gene intersection size > 20.

We then looked at results for glucose measurement (COLOC score = 3.6, **Figure 5C**), the trait with the greatest tissue expression difference between CVGs and RVGs. Individually, CVGs associated with glucose measurement were expressed in a median of 53 tissues, compared to 2 tissues for RVGs (**Figure 5A**). Driven by three genes - *G6PC2*, *SLC30A8*, and *KCNK16* - the set of RVGs was exclusively enriched for pancreas-specific expression (*p_adj_* = 4.0×10^−5^). *G6PC2* and *KCNK16* are highly constrained genes (LOEUF*_G6PC2_* = −0.17, LOEUF*_KCNK16_* −0.13), which may have contributed to the lack of significantly associated CVs identified in these genes. In contrast, *SLC30A8* is less constrained (LOEUF = 1.7) and was the singular gene shared between CVGs and RVGs for glucose measurement (**Figure 5C**). The remaining two RVGs, *ANKH* and *SFT2D3*, were not specifically expressed in relevant tissues and were not prioritized as important by the network colocalization analysis. Globally, the ubiquitously expressed CVGs and pancreas-specific RVGs in the glucose measurement network converged to crucial processes underlying glucose regulation (**Figure 5D**), such as AMPK signaling (*p_adj_* = 3.5×10^−29^), glucagon signaling (*p_adj_* = 2.2×10^−28^), insulin signaling (*p_adj_* = 1.5×10^−25^), and insulin resistance (*p_adj_* = 7.5×10^−24^). The glucose measurement network was also enriched for relevant human phenotypes, such as an increased waist-to-hip ratio (*p_adj_* = 7.5×10^−20^) and Type II Diabetes Mellitus (*p_adj_* = 1.4×10^−8^).

### Systems impacted by common and rare variants associated with neuropsychiatric traits

Using neuropsychiatric traits as an example, we examined the mechanisms underlying the concordance of common and rare variants. Among the 21 neuropsychiatric traits, 14 showed significant network convergence (*q* < 0.05, **Figure 6A**). Autism spectrum disorder (ASD) had the strongest convergence (COLOC score = 2.8), consistent with the high genetic heritability^22^ and the known contributions of both common and rare variants to this condition^12^. Given the reported genetic similarities across neuropsychiatric traits^23^, we assessed the similarity of the trait networks using the cosine similarity of network proximity scores for the union of the trait network genes. These similarities revealed two distinct clusters (**Figure 6B**). The larger cluster included ASD, bipolar disorder (BD), epilepsy, insomnia, schizophrenia (SCZ), and headache, while the smaller cluster included Alzheimer’s disease (AD), multiple sclerosis (MS), and obsessive-compulsive disorder (OCD). The greatest similarity was observed between ASD and insomnia, consistent with the high co-occurrence of these disorders^24^.

**Figure 6.**
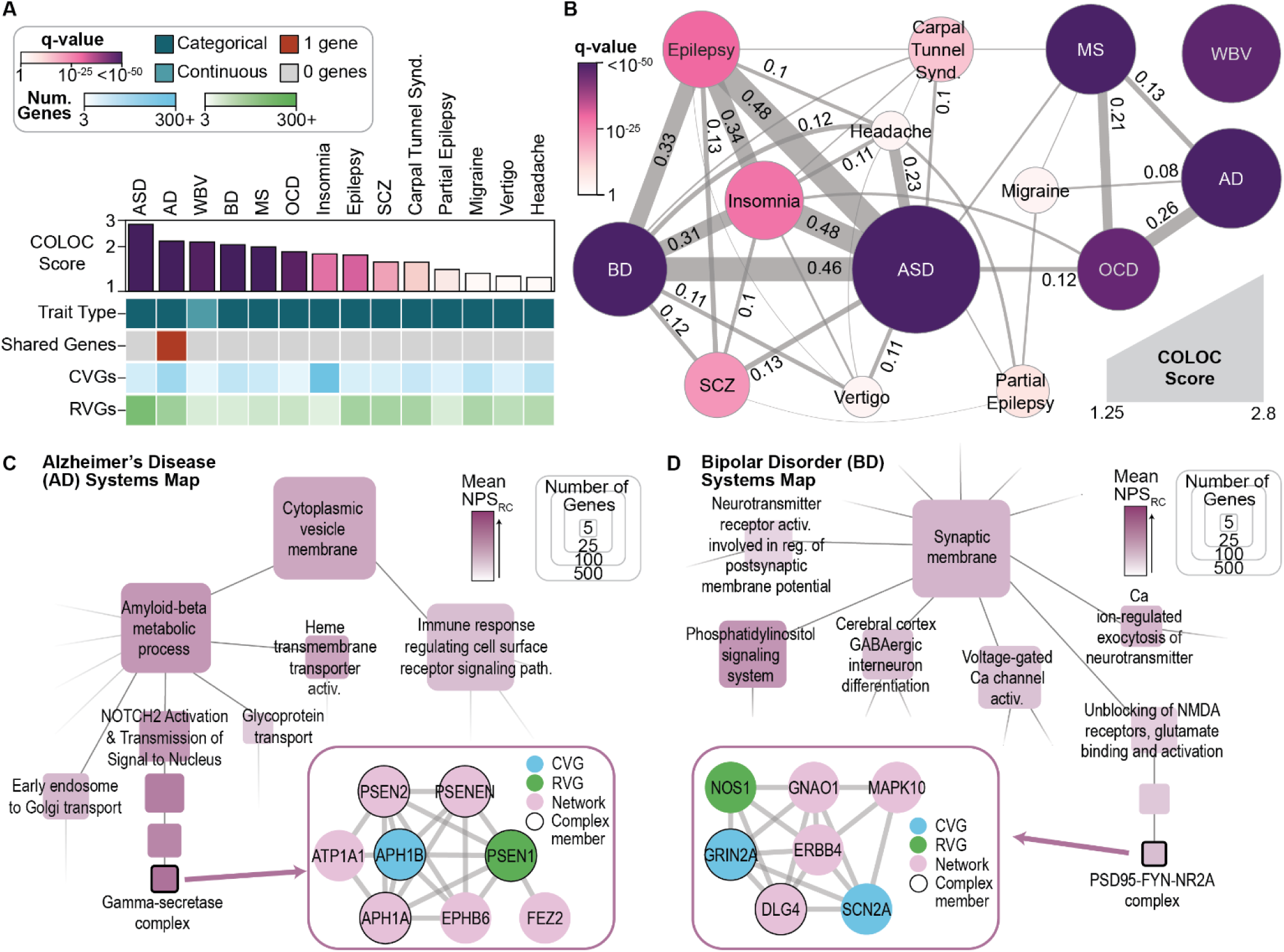
Systems impacted by common and rare variants for neuropsychiatric traits. **A)** Network colocalization results and trait features for the 14 neuropsychiatric traits with significant network convergence of CVGs and RVGs. **B)** Cosine similarity network of 14 neuropsychiatric traits. Edge weights and labels represent the cosine similarity of the individual trait networks. Edges shown for similarities ≥ 0.05 and edges labeled for similarities ≥ 0.10. Node size and color show the COLOC score and q-value, respectively. **C-D)** Subsets of the systems maps for Alzheimer’s disease (C) and bipolar disorder (D), generated via hierarchical community detection of the trait networks and annotated using g:Profiler (Methods). Node colors indicate the mean network proximity score (NPSRC) across all system genes, and node size indicates the number of system genes. Insets show the underlying network for selected systems derived from PCNet 2.0, with node colors reflecting the gene status as input associations (CVG, RVG) or prioritized by the network analysis (Network). ASD: Autism Spectrum Disorder, AD: Alzheimer’s Disease, BD: Bipolar Disorder, MS: Multiple Sclerosis, OCD: Obsessive-Compulsive Disorder, SCZ: Schizophrenia. WBV: While Brain Volume. See also Supplemental Figure 3.

Next, we examined the three traits with the most significant network convergence: AD, BD, and ASD. For AD, CVGs and RVGs converged on two major biological processes involved in AD pathogenesis (**Figure 6C**, **Supplemental Figure 3A**): one related to amyloid beta metabolism (*p* = 6.2×10^−21^) and the other to immune signaling (*p* = 1.1×10^−37^). One protein complex impacted by both CVs and RVs was the gamma-secretase complex, which catalyzes the proteolysis of the amyloid-beta precursor. RVs in the subunit gene presenilin 1 (*PSEN1*) were significantly associated with AD (*p_RV_*=3×10^−7^)^3^, as was the CV rs117618017 (*p_CV_* = 4×10^−20^)^25^, which falls within the gamma-secretase subunit gene *APH1B*. Therefore, despite implicating different genes within the complex, both CVs and RVs may influence AD via the gamma-secretase complex. For BD, CVGs and RVGs converged to a range of synaptic and neuronal signaling processes (**Figure 6D**, **Supplemental Figure 3B**), including phosphatidylinositol signaling (*p* = 1.2×10^−31^), GABAergic interneuron differentiation (*p* = 1.3×10^−5^), and the regulation of neurotransmitters such as glutamate (*p* = 9.4×10^−12^). Within glutamate signaling, CVs were identified in the glutamate ionotropic receptor subunit *GRIN2A* (*p_CV_* = 9×10^−11^)^26^ and the sodium voltage-gated channel subunit *SCN2A* (*p_CV_* = 2×10^−9^)^26^, alongside RVs in nitric oxide synthase 1 (*NOS1*, *p_RV_* = 4.1×10^−7^)^27^. In concert with other interacting proteins, perturbations to these proteins may collectively influence BD via impacts on the excitatory synapse.

Given the high network similarity observed between BD and ASD (**Figure 6B**), we compared the functions identified by convergent CVGs and RVGs in each disorder (**Figure 6D**, **Supplemental Figure 3C**). For both BD and ASD, the CVGs and RVGs converged broadly on synapse components. However, variants associated with ASD specifically implicated synapse organization (*p* = 4.1×10^−107^) and developmental processes such as netrin signaling (*p* = 1.3×10^−11^). On the other hand, variants associated with BD implicated systems related to metabolic compounds, such as methionine (*p* = 6.5×10^−5^) and high-density lipoprotein (*p* = 7.5×10^−6^). These findings underscored the shared neuronal mechanisms of BD and ASD, while also identifying distinct processes impacted by both common and rare variants in each disorder.

## DISCUSSION

In this study, we find that common variants (CVs) and rare variants (RVs) converge on shared biological networks for most complex human traits (76%, **Figures 2 & 3**), despite only a quarter of associated CVs and RVs impacting shared effector genes (**Figure 1**). Gene-level comparisons^6,8,28^ may thus underestimate the shared biological functions of common and rare variants. The degree of network convergence between CVs and RVs varied across traits, with greater concordance for traits that are quantitative, highly heritable, or involve more mutationally constrained genes (**Figure 4**, **Table 1**). The finding that traits with high SNP heritability exhibit stronger network convergence (**Figure 4A**) builds on previous findings that common and rare variant heritability are correlated^29^ and that incorporating rare variants alongside common variants enhances heritability prediction^5^.

Traits with RVs in tissue-specific genes also showed stronger CVG/RVG convergence (**Figure 4H**, **Figure 5**), suggesting that the focused expression of these genes may help define the core structure of the trait’s genetic architecture. Results from RV burden tests support this interpretation, as they tend to identify genes that are more trait-specific^7^. However, differences in the specificity of regulatory effects between variant classes have been reported: Li et al. reported that RV expression quantitative trait loci (eQTLs) are more tissue-specific than CV eQTLs^30^, whereas Cuomo et al. found that CV eQTLs are more cell-type-specific than RV eQTLs^31^. Future integration of eQTLs alongside common and rare variants could clarify the roles of these tissue-specific regulatory variants.

The trait networks generated from convergent CVGs and RVGs provide opportunities for multi-scale interrogation of how genetic variation converges on biological mechanisms. At a broad biological process or molecular pathway level, the influence of common and rare variants for Alzheimer’s disease (AD) was partitioned into two facets: amyloid-beta metabolism and immune signaling (**Figure 6C**). At the more granular level of individual protein complexes, we identified a convergence of AD-associated variants on gamma-secretase, a protease complex that has been studied as a potential drug target for this disease^32^. These results illustrate how common and rare variants, even when located in different genetic loci, implicate similar functions across multiple levels of biological organization. Furthermore, in bipolar disorder (BD) and autism spectrum disorder (ASD), genetic variants converged on synaptic structure and function (**Figure 6D**, **Supplemental Figure 3B-C**). However, they diverged in other processes, including methionine metabolism, consistent with reports that S-adenosyl-L-methionine (SAMe) can induce mania in BD^33^.

The network convergence results were robust across multiple study designs for the same trait (**Figure 3**), showing the benefit of a network approach in prioritizing biologically consistent genes from inputs containing noise^18^. To maximize the accuracy of the input associations, we examined CVs that fell directly within gene regions. However, while SNP genomic location remains foundational for identifying causal genes from CVs^34^, it cannot capture the full complexity of gene regulation. SNP-to-gene methods incorporating additional factors, such as linkage disequilibrium^35^ or functional annotations^34^, could improve the identification of causal genes; limited availability of summary statistics and the inclusion of mixed ancestry populations precluded their use in this study. Nonetheless, future studies could leverage refined methods for SNP-to-gene mapping and study selection to improve the accuracy of input genes.

In conclusion, by harmonizing studies of common and rare variants within a comprehensive biological knowledge network, we find that these studies implicate complementary genes and consistent biological systems. Our results provide a foundation for systematically understanding the shared roles of diverse genetic signals. Future applications of this framework could incorporate additional variant types, such as de novo mutations and copy number variants, to achieve a more complete understanding of the genetic etiology of human traits. To support further research, we have made molecular trait networks available for all 283 convergent traits via the Network Data Exchange (NDEx)^36^.

## DATA & CODE AVAILABILITY

All common and rare variant associations were sourced from the GWAS Catalog (https://www.ebi.ac.uk/gwas/) and RAVAR (http://www.ravar.bio). Associated study accession numbers are listed in **Supplemental Table 1**. Code used for data processing, analysis, and visualization is available on GitHub at https://github.com/sarah-n-wright/CARVA. Network resources produced in this study are available on the Network Data Exchange (NDEx, https://ndexbio.org) as part of the network set: <u>Common and rare genetic variants show network convergence for a majority of human traits</u>. DOIs to be issued at the time of publication.

## Data Availability

All data produced in the present work are contained in the manuscript

https://www.ebi.ac.uk/gwas/api/search/downloads/alternative

http://www.ravar.bio/api/download/static/gene_fulltable.txt

## ACKNOWLEDGEMENTS

This work was supported by the following grants from the National Institutes of Health: NIMH U01 MH115747 to T.I., and NIDA P50 DA037844 to T.I. This work was also supported by the following grants from the California Institute for Regenerative Medicine: ReMIND DISC4-16322 and ReMIND DISC4-16377.

## DECLARATION OF INTERESTS

T.I. is a co-founder, member of the advisory board, and has an equity interest in Data4Cure and Serinus Biosciences. T.I. is a consultant for and has an equity interest in Ideaya Biosciences and Eikon Therapeutics. The terms of these arrangements have been reviewed and approved by the University of California, San Diego, in accordance with its conflict-of-interest policies.

## METHODS

### Data Curation

#### Biological Knowledge Networks

All networks were sourced from the Network Data Exchange (NDEx, http://ndexbio.org/). PCNet 2.0 (https://doi.org/10.18119/N9JP5J) contains all interactions present in at least two of fifteen high-performing interaction networks^17^. PCNet 2.2 (https://doi.org/10.18119/N9960N) is a reduced version of PCNet 2.0 containing all interactions present in at least two of ten high-performing interaction networks without co-citation evidence^17^. We sourced standardized versions of the STRING v12.0^37^ network and the HumanNet v3 XC^38^ network from the ‘State of the Interactomes - Source Networks’ network set (https://doi.org/10.18119/N95C9J). The STRING network was filtered to interactions with a score > 700 to define the STRING High Confidence (HC) network.

#### Gene Identifier Mapping

Gene identifiers were mapped using the gene_mapper module from neteval v0.2.2^17^ (https://pypi.org/project/neteval/). Input identifiers were first updated to the most recent versions in the original database. Unless otherwise specified, all gene identifiers were converted to NCBI Gene IDs.

#### Trait Semantic Similarity

To assess the similarity of free-text trait descriptions, we embedded the trait descriptions using the pretrained Sentence-BERT (SBERT) via the sentence_transformers^39^ Python package. Following expansion of common acronyms (e.g., HDL: high-density lipoprotein), all descriptions were embedded using the model ‘all-MiniLM-L6-v2’. The cosine similarity between two trait embeddings was used to quantify the trait similarity. EFO term descriptions were sourced from the Experimental Factor Ontology (http://www.ebi.ac.uk/efo/efo.obo) via the obonet v1.0.0 (https://pypi.org/project/obonet/1.0.0/) Python package.

#### Common Variant Associations

Results from common-variant genome-wide association studies (GWAS) were downloaded from the GWAS Catalog on January 29, 2025 (All associations v1.0.2, https://www.ebi.ac.uk/gwas/api/search/downloads/alternative), along with accompanying trait (https://www.ebi.ac.uk/gwas/api/search/downloads/trait_mappings) and study (All studies v1.0.2.1, https://www.ebi.ac.uk/gwas/api/search/downloads/studies/v1.0.2.1) metadata. Gene-level associations were taken from the MAPPED_GENE column provided by the GWAS Catalog. Briefly, a SNP is mapped to a gene if it falls within the gene region, as defined by the Ensembl location^15^. We filtered associations to those falling within a single gene region to define a higher confidence set of associated genes. Where multiple SNP associations mapped to a single gene, the strongest p-value was maintained for the gene association. The set of common-variant associated genes (CVGs) for each trait was defined as the set of distinct genes tagged by SNPs with *p* < 1×10^−5^. Based on study metadata, studies were excluded if they: 1) listed a background trait (‘MAPPED BACKGROUND TRAIT’), 2) utilized targeted sequencing or exome sequencing (‘GENOTYPING TECHNOLOGY’) to ensure all studies represent genome-wide analyses, 3) did not list study population size (‘INITIAL SAMPLE SIZE’), or 4) reported fewer than three distinct gene associations for genes present in the PCNet2.0 network at *p* < 1×10^−5^.

Mappings between reported traits (‘DISEASE/TRAIT’) and Experimental Factor Ontology (EFO) terms (‘MAPPED_TRAIT_URI’) in the GWAS Catalog are not one-to-one. Therefore, we standardized EFO trait mappings by: 1) maintaining all one-to-one mappings, 2) excluding studies listing multiple EFO terms for a single reported trait, and 3) taking the EFO terms with the highest semantic similarity for the same reported trait mapped to multiple EFO terms across different studies.

#### Rare Variant Associations

We sourced rare variant gene-level association results and related metadata from the Rare Variant Association Repository (RAVAR) database^14^ on June 11, 2024 (http://www.ravar.bio/api/download/static/gene_fulltable.txt). Additional metadata, including population size, study design, and population cohort, were manually curated from the associated publications for the included studies. Where available, we took the population sample size for individual traits within multi-trait studies. The set of rare-variant associated genes (RVGs) for each trait was defined as the set of distinct genes with gene-level association *p* < 1×10^−4^. Studies were excluded if: 1) they collated associations from literature reviews or sequencing studies that were not genome-wide or exome-wide, 2) population size could not be determined, 3) they reported fewer than three distinct gene associations for genes present in PCNet2.0 at *p* < 1×10^−4^

Mappings of reported trait descriptions (‘Reported Trait’) and EFO terms were harmonized with the GWAS catalog, using the same procedure described for the common variant studies. If a reported trait did not have an exact match in the GWAS catalog, the EFO mapping supplied by RAVAR (‘Trait Ontology id’) was maintained. This process ensured consistent EFO mappings between the GWAS Catalog and RAVAR.

#### Common and Rare Trait & Study Matching

We identified traits with matched common and rare variant results using the mapped EFO terms. For traits with multiple independent studies, we prioritized a single study for the initial analysis and up to four additional studies for replication analyses.

The initial common variant study per trait was selected as the GWAS with the largest population sample size, after filtering for high trait similarity of reported trait description (‘DISEASE/TRAIT’) with mapped EFO term description and availability of cohort information and summary statistics. The initial rare variant study per trait was selected as the study with the largest population sample size, after filtering for high trait similarity between the reported trait description (‘Reported Trait’) and the mapped EFO term description, and the availability of cohort information. After matching based on EFO terms, the trait semantic similarities of matched studies (GWAS Catalog: ‘DISEASE/TRAIT’, RAVAR: ‘Reported Trait’) were calculated. All study combinations with a similarity of less than 0.7 were assessed manually, and only those that represented synonyms of the same trait were maintained for the analysis.

Up to four additional GWAS and four additional rare variant studies were selected per trait for replication analysis. Studies were required to have a cosine similarity between the reported trait and mapped EFO term of at least 0.7. Where possible, studies utilizing different population cohorts were prioritized, with further prioritization based on the largest population sample sizes.

Information on all selected studies and accompanying gene sets can be found in **Supplemental Table 1**. For each combination of common and rare studies within each trait, the gene-level convergence was calculated by a hypergeometric test between the sets of CVGs and RVGs. A background of 19000 was used to approximate the total number of human protein-coding genes.

#### Trait and Study Properties

Traits were manually classified as categorical if they involved a case-control study or other discrete phenotype, or continuous if they involved a quantitative measurement. All traits were manually assigned to high-level biological domains. Domains with fewer than 5 traits in the analysis set were assigned to the ‘Other’ category for visualization purposes.

Trait SNP heritability estimates were sourced from the Neale Lab Round 2 analysis^20^, calculated via partitioned LD Score regression (LDSC) for 4236 phenotypes in the UK Biobank. We excluded ten pigmentation and bilirubin phenotypes based on their unusually high standard error (SE) and low polygenicity, as flagged in the UKB heritability analysis. We also excluded three traits flagged as having ordinal coding issues based on the column ‘isBadOrdinal’. For each of our 373 traits, we identified the most semantically similar trait in the UKB heritability results and manually curated the matches to remove duplicates and imprecise matches. In total, we identified heritability estimates for 197 traits.

Disease prevalence estimates were sourced from the Global Burden of Disease (GBD) study from the Institute for Health Metrics and Evaluation^21^. We extracted disease prevalence as a percent of the population for the five years from 2017 to 2021, including all locations, ages, and sexes. We took the mean prevalence across the five-year period. For each of our 373 traits, we identified the most semantically similar trait in the GBD prevalence results and manually curated the matches to remove duplicates and imprecise matches. In total, we identified prevalence estimates for 87 traits.

#### Gene Properties and Annotations

Gene mutational constraint estimates were sourced from gnomAD v4.1^16^ (https://gnomad.broadinstitute.org/data, Constraint metrics TSV). For each gene, we extracted the LOEUF (loss-of-function observed/expected upper bound fraction)^16,40^ and the synonymous intolerance Z-score^40,41^. The LOEUF metric provides a continuous metric of a gene’s intolerance to loss-of-function variants, with a threshold of LOEUF < 0.6 recommended for defining LoF-constrained genes. For each distinct gene symbol, we prioritized entries associated with canonical and MANE Select transcripts, followed by prioritization of entries with an NCBI Gene ID assigned. Gene region annotations were sourced from Ensembl v113^15^, downloaded on February 14, 2025. Entries were filtered to transcripts on hg38 primary chromosome assemblies with a mapped HGNC Symbol. Gene length was calculated as the difference between the start (bp) and end (bp) positions. Genes with conflicting entries were excluded. Median gene-level mRNA expression by tissue was sourced from GTEx v8^42^. For genes with multiple transcripts, we consolidated the values for all transcripts within each tissue. We took the mean of all transcripts if all values had a similar magnitude (all TPM observations above or below 10^−4^); otherwise, we took the maximum. Overall expression levels were calculated as the mean across all tissues. The number of expressed tissues per gene was defined as the number of tissues with TPM > 1. Where a gene had multiple transcripts, the transcript with the highest mean expression was selected. Functional Gene Ontology (GO) associations were downloaded from NCBI using goatools^43^ on November 25, 2024. Entries with the negating qualifier ‘NOT’ were excluded. We extracted the number of distinct GO IDs associated with each human NCBI Gene ID across the three branches of GO: Cellular Component (CC), Biological Process (BP), and Molecular Function (MF). Missing genes were assigned a value of zero.

Summary values were calculated for each set of RVGs and CVGs by taking the median value for all genes in the gene set. Genes with missing values were excluded. Paired differences between common and rare gene sets for matched traits were assessed by the Wilcoxon Signed-Rank Test with BH correction.

### Quantitative Network Colocalization (Q-NetColoc)

#### Network Proximity Scores (NPS)

For network propagation and colocalization, we used the Python package NetColoc v0.1.8^18,19^ (https://pypi.org/project/netcoloc/). First, sets of common and rare variant associated genes (CVGs and RVGs) were propagated through the network using a Random Walk with Restart process^44^. For efficient computation, NetColoc utilizes the closed-form solution for the random walk process described by Equation 1:

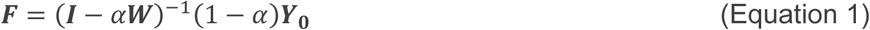

Where ***F*** is the stable heat vector for all nodes, ***Y***_0_is the initial heat for all nodes, ***W*** is the degree-normalized adjacency matrix of the undirected network, and α ∈ (0, 1) is the dissipation constant. For the original NetColoc method, the initial heat vector ***Y***_0_is assigned 1 for all genes in the input set (all genes with association p-value below a threshold θ), and 0 otherwise. We refer to this as binary NetColoc (B-NetColoc). Given the lenient thresholds used to define CVGs and RVGs, we modified NetColoc to weight all CVGs and RVGs based on the negative-log_10_ association p-value to appropriately up-weight the most confident associations. Therefore, we defined quantitative NetColoc (Q-NetColoc), where ***Y***_0_is assigned the value of −*log*_10_(*p*) for each associated gene with *p* < θ, and 0 otherwise. Values in ***Y***_0_were normalized such that the sum of all values was 1.

Following network propagation, network proximity scores (NPS) were calculated for each gene in the network as a Z-score of the observed heat relative to a background null distribution. This null distribution was formed for each gene via network propagation of degree-matched random gene sets. First, all network genes were binned by network degree to create bins containing a minimum of *β* genes. For B-NetColoc, we randomly selected gene sets of equal size from the bins corresponding to the input gene set. For Q-NetColoc, we shuffled the heats of all genes within each bin in order to maintain the degree-heat correlation. For each gene *g*, we then calculated NPS as:

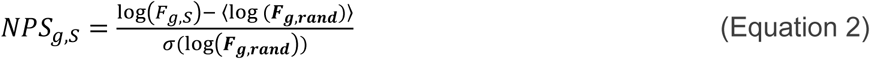

Where *F*_*g*,*s*_is the observed heat at *g* after propagation of the gene set *S*, and ***F***_*g*,_*_rand_* is the null distribution of heats from randomized gene sets. The Mean of a vector is denoted by 〈 〉 and σ denotes the standard deviation of a vector. All values are log-transformed to ensure the distributions are approximately normally distributed. Following the calculation of NPS_C_and NPS_R_for all genes, we calculated the combined network proximity NPS_CR_as the product of the independent gene scores:

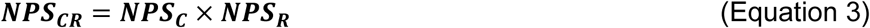

#### Colocalized Trait Networks and COLOC Score

To assess the convergence of CVGs and RVGs within the network, we defined the colocalized trait network *G*_trait_ as:

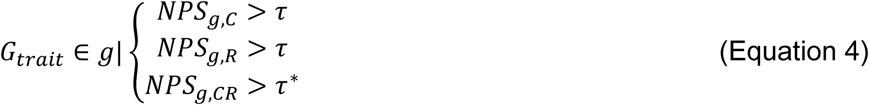

Where τ and τ^∗^ are thresholds on NPS. The observed size of each *G*_trait_ was compared to a permuted null distribution. We permuted the labels of NPS_C_1000 times, each time calculating the number of genes meeting the thresholds for inclusion in the trait network (Equation 4). For genes shared between the common and rare input sets, labels were permuted separately to maintain the higher expected distribution for these genes. The significance of network colocalization (COLOC P) was defined by a Z-test comparing the observed size of *G*_trait_to the expected size of *G*_trait_. We defined the observed over expected size of the trait network as the network colocalization (COLOC) score.

#### Optimization and Benchmarking of Q-NetColoc

Our implementation of Q-NetColoc was benchmarked and optimized using GWAS results from the GWAS Catalog for a random sample of traits not included in the main analysis. In addition to the criteria outlined in <u>Common Variant Associations</u>, we selected studies with publicly available summary statistics, a single-ancestry cohort, and a defined genome build. In total, we generated a testing set comprising 87 studies covering 43 distinct traits (**Supplemental Table 5**). From each study, we extracted the common variant-associated genes (CVGs) as described in <u>Common Variant Associations</u>. All sets of CVGs were partitioned into two non-overlapping sets of equal size. Test pairs were formed by matching two halves of the same study, and control pairs were formed by matching two halves of studies of unrelated traits. For Q-NetColoc, all p-values below the association p-value threshold (θ) were transformed to negative-log_10_ scores and normalized by dividing by the sum of all values in the set. For B-NetColoc, p-values were binarized as significant (1) and non-significant (0) based on the association p-value threshold (θ).

##### Assessment Metrics

We assessed the performance of different NetColoc configurations using the Area Under the Receiver Operating Characteristic curve (AUROC). Within each group of configurations below, we subset the results to all pairs evaluated by those configurations and a matching number of control pairs. AUROC was calculated using the roc_auc_score function from scikit-learn^45^. AUROC was calculated for the COLOC score and the associated colocalization p-value (COLOC P).

##### Thresholds

We first assessed the optimal thresholds τ and τ^∗^ for defining the trait network (Equation 4) for Q-NetColoc and B-NetColoc. Thresholds were scanned in the range {1.0, 1.5, 2.0, 2.5, 3.0, 3.5, 4.0} for τ^∗^ ≥ τ. A default bin size of *β* = 10 for degree-matched randomization was used. The parameter τ had a greater effect on the observed AUROC than τ^∗^, with lower values giving better performance for Q-NetColoc and B-NetColoc (**Supplemental Figure 4A**). Therefore, we proceeded with τ = 1, paired with a stricter value of τ^∗^ = 3, for all subsequent optimization and analysis tasks.

##### Bin Size

The bin size (*β*) for performing degree-matched randomization balances the randomization of scores with the maintenance of the degree-score correlation. We tested values of the bin size parameter *β* ∈ {5, 10, 20, 40, 80, 160} for B-NetColoc and Q-NetColoc. Based on the AUROC values, we determined that the optimal bin size was *β* = 20 (**Supplemental Figure 4B**).

##### Association P-value

Initial testing was performed using a lenient association p-value threshold of θ*_lenient_* = 1 × 10^−5^ for determining CVGs. While this allows for the inclusion of more genes and traits, it also increases the inclusion of false-positive associations. Therefore, we also tested a strict threshold of θ*_strict_* = 1 × 10^−8^. The lenient threshold consistently provided better separation between the test and control pairs (**Supplemental Figure 4C**). For example, looking at effect sizes from Q-NetColoc, the lenient threshold gave AUROC=0.84, compared to AUROC=0.82 for the strict threshold. Across all tests, minimal differences were observed between Q-NetColoc and B-NetColoc. Therefore, we determined that the benefits of including the appropriately weighted sub-threshold associations outweighed the potential inclusion of more false positives.

##### SNP-to-Gene Mapping

Finally, we compared the default SNP-to-Gene mapping approach (S2G) to the gene aggregation approach MAGMA^35^. For each testing study, we standardized the summary statistics, annotated the SNPs using the assigned genome build, and calculated gene-level p-values using MAGMA v1.10 with the most relevant population reference from the 1000 Genomes Project^46^, based on the listed study ancestry. MAGMA and all auxiliary files were downloaded from https://cncr.nl/research/magma/ on December 5, 2024. For MAGMA results, we defined the strict threshold as θ*_strict_* = 2.5 × 10^−6^, corresponding to the typical multi-hypothesis correction for the number of coding genes. We defined the lenient threshold as θ*_lenient_*= 1 × 10^−4^. Overall, the MAGMA results provided slightly better separation between test and control pairs. For example, based on the COLOC score, MAGMA achieved an AUROC=0.89 (**Supplemental Figure 4D**). As with the S2G gene sets, the lenient threshold yielded better results for MAGMA. Therefore, where practical, MAGMA gene-level association statistics can boost NetColoc performance. However, variability in the structure and availability of summary statistics and population ancestry precluded the use of MAGMA for our primary analysis.

#### Network Colocalization of Matched CVGs and RVGs using Q-NetColoc

For the main analysis, Q-NetColoc was implemented with a bin size *β* = 20, trait network thresholds of τ = 1 and τ^∗^ = 3, and PCNet2.0 as the underlying network. Input genes were weighted based on the negative-log_10_ association p-value, with association p-value thresholds of θ_CVG_ = 1 × 10^−5^ for CVGs and θ_RVG_ = 1 × 10^−4^ for RVGs.

The initial network colocalization was performed using the top common variant study and the top rare variant study per trait, as defined in <u>Common and Rare Trait & Study Matching</u>. The additional analyses were then performed with all pair-wise combinations of common variant and rare variant studies per trait. To avoid redundant calculations, NPS were calculated once for each set of CVGs or RVGs and reused across different study combinations. Multiple hypothesis correction was performed on the network colocalization p-values using a BH correction. Prioritized input genes were defined as input CVGs achieving NPS_*g*,R_ ≥ 1, and input RVGs achieving NPS_*g*,C_ ≥ 1. For each trait, the optimal combination of common variant study and rare variant study was defined as the study combination achieving the highest COLOC score. Using the 373 optimal study combinations, Q-NetColoc was implemented using PCNet2.2, HumanNet v3, and STRING-HC as the underlying network, with all other parameters equal. Results were generated for each network for all traits with at least three CVGs and at least three RVGs in that network.

### Analysis and Interpretation of Common-Rare Colocalized Trait Networks

#### Enrichment for Tissue-Specific Genes

Tissue-specific enrichment analysis (TSEA) was performed using the R package TissueEnrich^47^ for traits with significant network colocalization. For each trait, we defined four gene sets: all genes in the colocalized trait network, prioritized genes (input genes in the trait network), all input genes (the union of CVGs and RVGs), and shared input genes (the intersection of CVGs and RVGs). All genes were mapped to Ensembl gene identifiers. Tissue-specific genes were defined from RNA-seq results from the Human Protein Atlas (HPA^48^) data included in TissueEnrich using default parameters of fold change ≥ 5, minimum expression ≥ 1, and number of expressed tissues ≤ 7. For enrichment analysis, all genes classified as Tissue-Enriched, Tissue-Enhanced, or Group-Enriched were considered. The set of all genes in PCNet2.0, converted to Ensembl gene identifiers, was used as the background gene list, and multiple-hypothesis correction was performed using BH-correction. To calculate enrichment in trait-relevant tissues, we manually assigned relevant tissues based on the biological domain of each trait (e.g., Renal: Kidney). Where relevant tissues could not be clearly defined, traits were excluded from the analysis (e.g., anthropometric traits such as height and BMI).

TSEA and individual gene expression distributions for CVGs and RVGs associated with glucose metabolism were sourced from the TissueEnrich browser implementation using HPA (https://tissueenrich.gdcb.iastate.edu/), utilizing gene symbols for the trait-associated genes and background PCNet 2.0 genes.

#### Gene Set Features

We collected study design features and summary features of the genes in each set of CVGs (C) and RVGs (R) from all analyzed pairs of initial and additional studies (n = 1634 study pairs). For each study pair, we compiled the study population sizes (N_C_,N_R_) and the number of genes in the sets (*g*_C_, *g*_R_). For each pair of CVGs and RVGs, we calculated the Jaccard similarity (*J*_CR_) and identified the subnetworks of PCNet2.0 induced by the CVGs, the RVGs, and the union of CVGs and RVGs. We calculated the subnetwork densities ρ_C_ and ρ_R_ based on the number of edges between CVGs and between RVGs, respectively. We calculated the assortativity of CVGs and RVGs in the union subnetwork using the networkx^49^ function attribute_assortativity_coefficient. For gene set pairs with *J*_CR_ > 0, we took the mean assortativity of all common genes to disjoint rare genes and all rare genes to disjoint common genes: *A*_CR_ = (*A*_C,R−C_ + *A*_R,C−R_)/2.

For each of the gene properties LOEUF, gene size (GeneSize), mRNA expression (mRNA), number of expressed tissues (nTissues), and number of GO annotations (GO) described in *<u>Data Curation > Gene</u> <u>Properties and Annotations</u>*, we calculated the weighted mean value μ_CR_ = (*g*_C_μ_C_ + *g*_R_μ_R_)/ (*g*_C_ + *g*_R_) across CVGs and RVGs and the normalized difference in means (δ) defined as δ_CR_ = 2(μ_C_ − μ_R_)/(μ_C_ + μ_R_^)^ (**Supplemental Figure 2B**). Five trait study pairs with missing values for LOEUF were excluded to give a final sample size of 1629.

We transformed and normalized the features using scikit-learn^45^ based on the observed distributions. Features N_C_, N_R_, *A*_RC_, μ_LOEUF_, μ_Genesize_, μ_mRNA_, μ_GO_, δ*_LOEUF_*, δ*_Genesize_*, δ*_mRNA_*, δ*GO*, δ*_nTissues_* were quantile-normalized to a Gaussian distribution using QuantileTransformer. The features *g*_C_, *g*_R_, ρ_C_, ρ_R_, *J*_RC_, μ_nTissues_ were power-transformed using PowerTransformer. Finally, we added a one-hot encoded feature to indicate categorical traits (‘binary’) and a one-hot encoded feature to identify the study pairs with *J*_RC_ ^=^ 0 (‘jaccard_zero’). To ensure comparable distributions for all features, we then rescaled all features using StandardScaler.

#### Elastic Net Regression Analysis

To estimate the contribution of each feature to the observed network colocalization (COLOC score), we performed a regression analysis using Elastic Net. This model was chosen to balance feature selection and account for correlations between the input features (**Supplemental Figure 2A**). The target variable, COLOC score, was quantile-normalized using QuantileTransfomer with the number of quantiles set to the sample size. To determine the optimal penalty parameter (α) and L1-ratio, we performed a five-fold cross-validation analysis to select the parameters minimizing out-of-fold mean-squared error (MSE), using ElasticNetCV from scikit-learn^45^. We utilized a log-spaced grid of 20 α values between 10^−5^ and 10^1^, and L1-ratios in {0.1, 0.25, 0.4, 0.5, 0.6, 0.7, 0.8, 0.9, 0.95, 0.99, 1}, with all other parameters set to their defaults. Following cross-validation, we identified an optimal α of 0.014, and L1-ratio of 0.1. The variance explained by the optimized model was calculated using explained_variance_score from scikit-learn. The optimized model was used to calculate the residual COLOC score as the difference between the predicted and actual COLOC scores.

We then performed bootstrapping to estimate the confidence of the coefficient fits. We generated 2000 bootstrap replicates using sampling with replacement and fit Elastic Net to each replicate. Coefficients for each feature are reported as bootstrap means with corresponding 95% confidence intervals. Empirical p-values were calculated as:

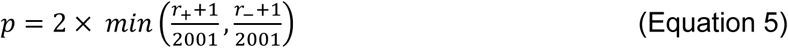

Where *r*_+_ is the number of coefficient estimates > 0 and *r*_−_ is the number of coefficient estimates < 0, followed by Bonferroni correction with the number of features.

#### Similarity of Trait Networks

The similarity of pairs of colocalized trait networks (*G_trait1_*: *G_trait2_*) was measured using cosine similarity. First, we identified the union of genes in both networks. Then we took the cosine similarity of common-rare network proximity scores (*NPS*_CR_) for genes in the union.

#### Construction of Hierarchical Systems Maps for Neuropsychiatric Traits

To identify the neurological systems underlying the convergence of CVGs and RVGs in neuropsychiatric disorders, we filtered the trait networks for Alzheimer’s disease (AD), bipolar disorder (BD), and autism spectrum disorder (ASD) to include only genes expressed in the brain. Genes were maintained if they had TPM > 1 in at least one of the thirteen brain regions of the Human Protein Atlas^50^ (HPA) brain region data version 24.0 (https://www.proteinatlas.org/about/download, rna_brain_region_hpa.tsv.zip), dated October 19, 2024.

Systems maps were constructed using the hierarchical community detection framework HiDeF^51^ via the Cytoscape package CyCommunityDetection v1.12.1^52^. We set the maximum resolution to 10, with all other parameters set to their default values. All network visualizations were constructed in Cytoscape^53^ v3.10.3 and uploaded to the Network Data Exchange^36^ (NDEx) as part of a network set (<u>Common and</u> <u>rare genetic variants show network convergence for a majority of human traits</u>). For interactive exploration of the systems maps and their underlying protein networks, we formatted the networks to comply with the hierarchical network schema (HCX, https://cytoscape.org/cx/cx2/hcx-specification) for CX2, using the NDEx2 Python Client v3.9.0 (https://github.com/ndexbio/ndex2-client). Briefly, the hierarchy network and parent trait network were imported in CX2 format. Then, the genes in each community were linked to the corresponding genes in the parent network. The uuid identifier of the parent network is then assigned as an attribute of the HCX hierarchy network. Finally, the HCX network is uploaded to NDEx, allowing access within Cytoscape Web^54^ (https://web.cytoscape.org). In Cytoscape Web, specific communities can be selected, and the corresponding protein subnetwork of PCNet2.0 will be dynamically displayed.

### Functional Enrichment and Annotation of Trait Networks and Hierarchical Systems Maps

Functional enrichments for the glucose measurement trait network were calculated using the g:Profiler^55^ website (https://biit.cs.ut.ee/gprofiler/gost) for KEGG Pathways and Human Phenotypes. Analysis was performed using human NCBI Gene IDs, the set of annotated genes as the background, and the BH correction for multiple-hypothesis testing.

For the AD, BD, and ASD systems maps, all communities were annotated using g:Profiler via CyCommunityDetection v1.12.1^52^ in Cytoscape. Enrichments were considered across a range of functional databases, including Gene Ontology, Reactome, CORUM, and WikiPathways. We set the maximum gene set size to 2000, the maximum p-value to 1×10^−4^, and the minimum overlap to 0.05. For visualization purposes, we retained annotated communities with at least five genes. Communities without a significant enrichment were labeled NA.

## SUPPLEMENTAL TABLE CAPTIONS

**Supplemental Table 1. All input common and rare studies, properties, and gene sets.** For common variant studies, the ‘Study Identifier’ is the GWAS Catalog accession number, the ‘Reported Trait’ is the original DISEASE/TRAIT listed for each study. For rare variant studies, the ‘Study Identifier’ is the PMID listed in RAVAR, and the ‘Reported Trait’ refers to the column of the same name in RAVAR. ‘Analysis Set’ refers to whether a study was used in the initial analysis of one study pair per trait, or as one of the additional studies used for replication analysis. ‘Mapped EFO’ terms include terms from the Experimental Factor Ontology (EFO), Mondo Disease Ontology (MONDO), Human Phenotype Ontology (HP), Gene Ontology (GO), and Ontology of Biological Attributes (OBA).

**Supplemental Table 2. All gene-level overlap and network colocalization results.** Each trait is defined by the mapped Experimental Factory Ontology term (EFO) and description (Trait). Common and rare variant studies used to generate the CVGs and RVGs are identified by GWAS Catalog accession numbers and PMIDs, respectively. ‘Analysis Set’ identifies the study combinations used in the initial and additional analyses. The number of genes identified as CVGs and RVGs are given by ‘nCommon’ and ‘nRare’, along with the size of the CVG-RVG intersection (nShared) and the associated overlap p-value ‘pShared’ (hypergeometric test). Results generated from Q-NetColoc with PCNet2.0 are provided: ‘Observed Size’ and mean ‘Expected Size’ of the trait network, as well as ‘Log2SizeOE’ and ‘COLOC Score’ based on the ratio of observed/expected size of the trait network. ‘COLOC p’ and ‘COLOC -logp’ are calculated from a Z-test comparing the observed size to the distribution of expected sizes. ‘Optimal COLOC’ identifies the study combination with the highest COLOC score for each trait. Additional Q-NetColoc analysis results with alternative networks are indicated by suffixes: PCNet2.2 (PC22), HumanNet v3 (HN), and STRING-HC (ST).

**Supplemental Table 3. SNP heritability and disease prevalence estimates**. Trait SNP heritability estimates were sourced from the Neale Lab Heritability browser. Observed h^2^ estimates are reported. Disease prevalence estimates were sourced from the Global Burden of Disease (GBD) study and averaged across the five-year period from 2017 to 2021.

**Supplemental Table 4. Trait and gene set features for all common-rare study combinations.** All initial and additional combinations of common and rare variant studies used to generate the CVGs and RVGs are identified by GWAS Catalog accession numbers and PMIDs, respectively. Features include the log2-transformed COLOC score, categorical trait status (binary), study population sample size (N), number of associated genes (g), Jaccard similarity of CVGs and RVGs (J_RC), flag for J_RC=0 (jaccard_zero), subnetwork densities (density), subnetwork assortativity (assortativity_RC), mutational constraint (LOEUF), gene length (GeneSize), number of associated GO terms (GO), mean mRNA expression (mRNA), and number of tissues with expression > 1 TPM (nTissues). Features with the prefix ‘Mean_’ represent the mean feature across CVGs and RVGs, and features with the prefix ‘DiffRC_’ represent the normalized difference in mean feature value between CVGs and RVGs.

**Supplemental Table 5. NetColoc testing results.** Results from the testing of Binary (B-NetColoc) and Quantitative (Q-NetColoc) implementations of network colocalization. Trait pairs were assigned to Test if they represented the same study, and Control if they represented unrelated studies. Run parameters utilized are given by: Bin Size - the degree-normalization bin size (*β*) used for calculation of network proximity scores (NPS), T & T* - the thresholds on NPSR and NPSC (τ) and NPSRC (τ^∗^) for defining the colocalized subnetwork, LenientOrStrict - the association p-value threshold (θ) for identifying CVGs and RVGs, and SNP-to-Gene - the method used to map CVs to CVGs. ‘Parameter Tested’ indicates the analysis group that results were used in: bin size testing (bin), colocalized network threshold testing (th), or SNP-to-Gene mapping and association p-value testing (GeneMappingAndAssocP). Results generated from NetColoc with PCNet2.0 are provided: ‘Observed Size’ and mean ‘Expected Size’ of the trait network, ‘Log2COLOC score’ and ‘COLOC p’.

## SUPPLEMENTAL FIGURES

**Supplemental Figure 1.**
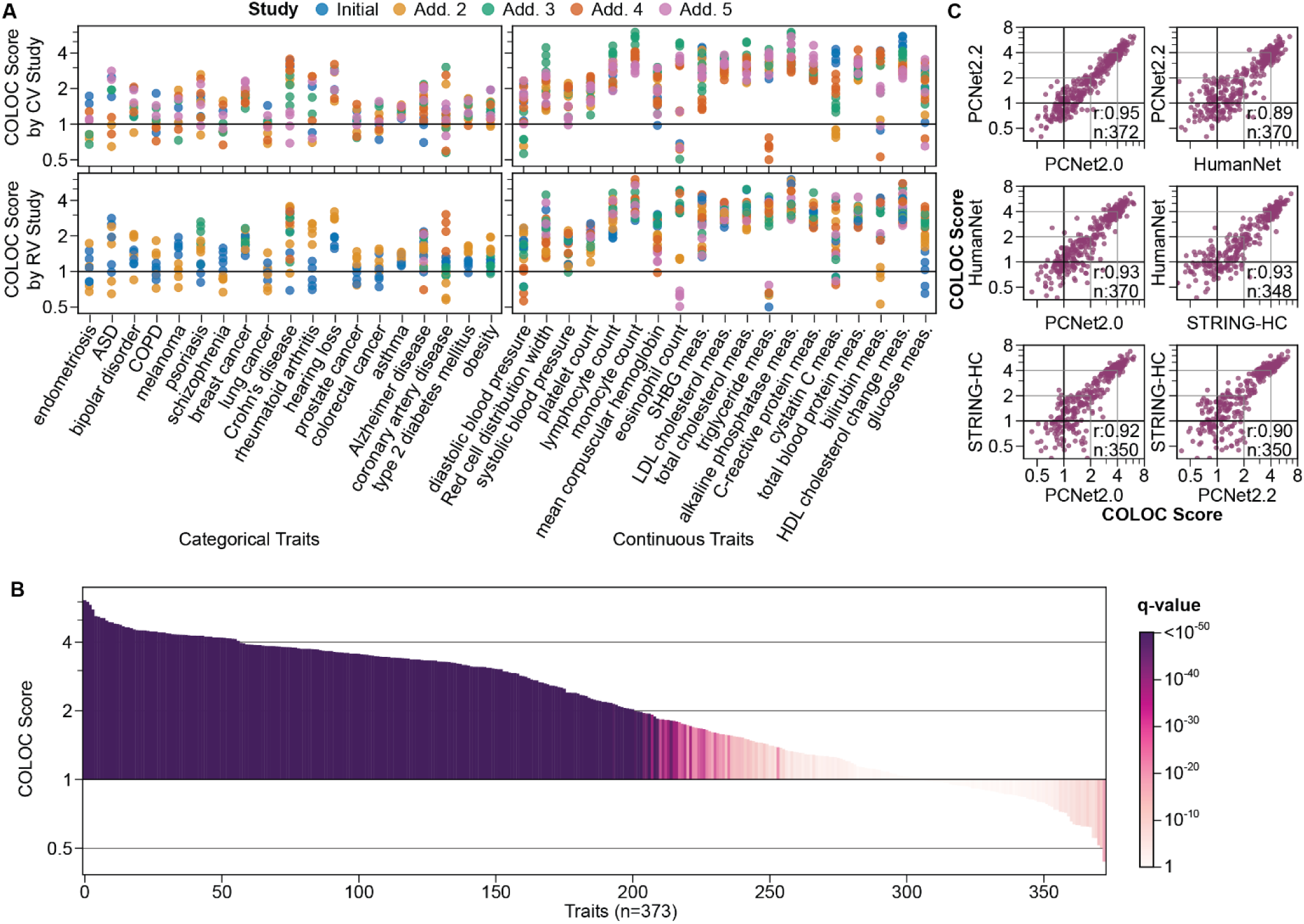
Robustness of the network convergence results across different studies and networks. **A)** COLOC scores for categorical traits with ≥ 10 rare-common study combinations (left, n=19) and continuous traits with ≥ 20 rare-common study combinations (right, n=19). The top plots are colored by common study, and the bottom plots are colored by rare study. **B)** Network colocalization scores for 373 traits after taking the highest COLOC score from all available rare-common study analyses per trait. Traits are ordered by COLOC score, and bar color indicates q-value of the COLOC score based on 1000 permutations with BH-correction. **C)** Pairwise comparison of COLOC scores across four biological networks. Correlation (r) calculated using Spearman correlation. The number of traits that could be assessed by both networks is given by n. Related to Figure 3.

**Supplemental Figure 2.**
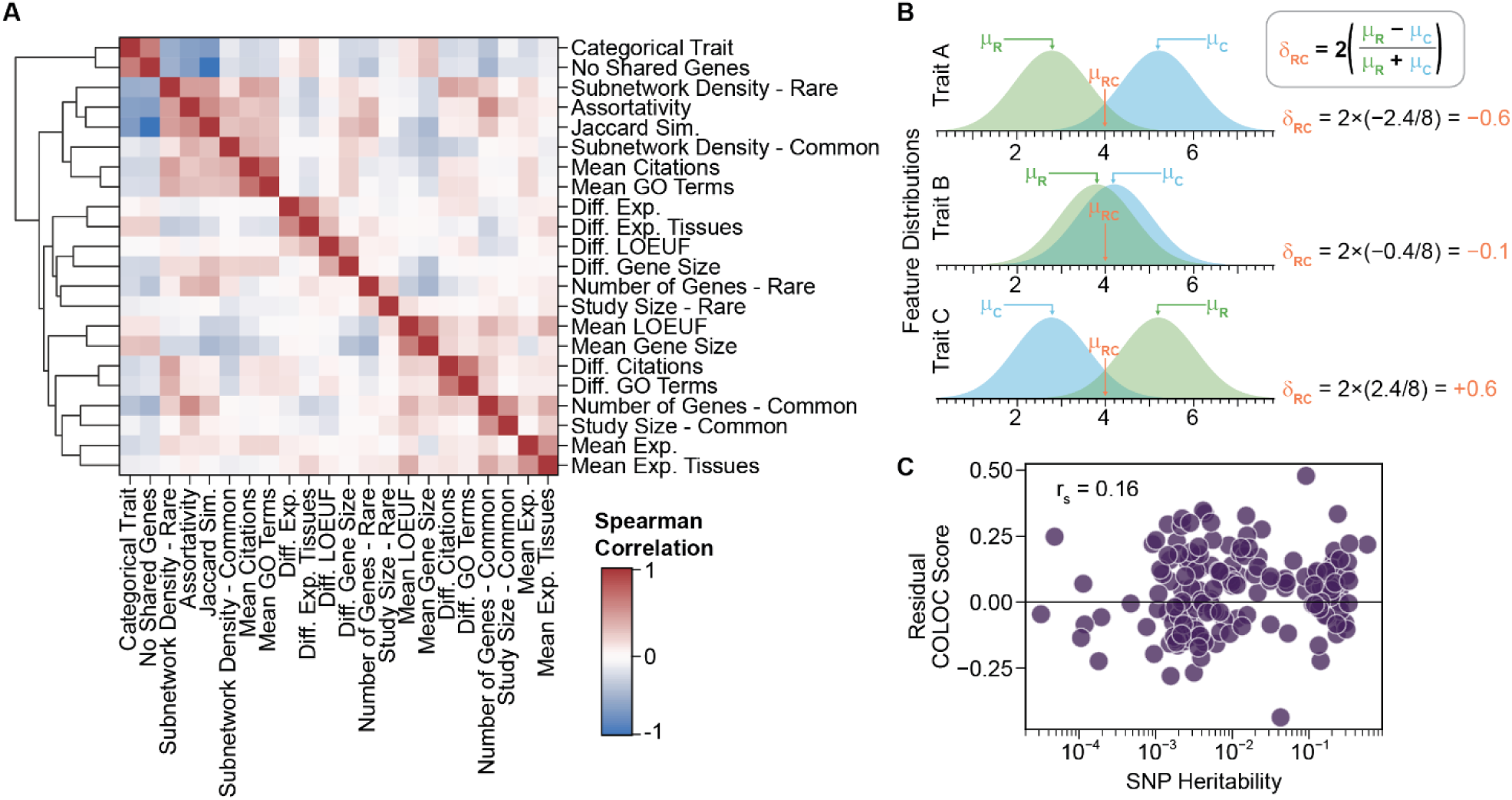
Expanded analysis of gene and trait features. **A)** Correlation of gene and trait features over 1634 common-rare study combinations for 373 traits. Jaccard similarity was calculated between CVGs and RVGs. Gene features were summarized by the overall mean of CVGs and RVGS, and the difference between CVGs and RVGs. Difference features were calculated as in panel B. LOEUF: Loss-of-function intolerance observed/expected upper fraction (mutational constraint). Exp.: median mRNA expression across all tissues. Exp. Tissues: Number of tissues with mRNA expression > 1 TPM. **B)** Schematic showing the calculation of mean (μ) and difference (δ) metrics for gene features. Distributions represent the hypothetical feature values for all CVGs (blue) and RVGs (green) of a given trait. Where the RVGs have higher average property values, the value (δ) will be positive. **C)** Residual COLOC score as a function of the SNP Heritability. The residual COLOC scores were calculated as the difference between the observed COLOC score and the COLOC score predicted by the Elastic Net regression model following five-fold cross-validation. Spearman Correlation reported. Related to Figure 4.

**Supplemental Figure 3.**
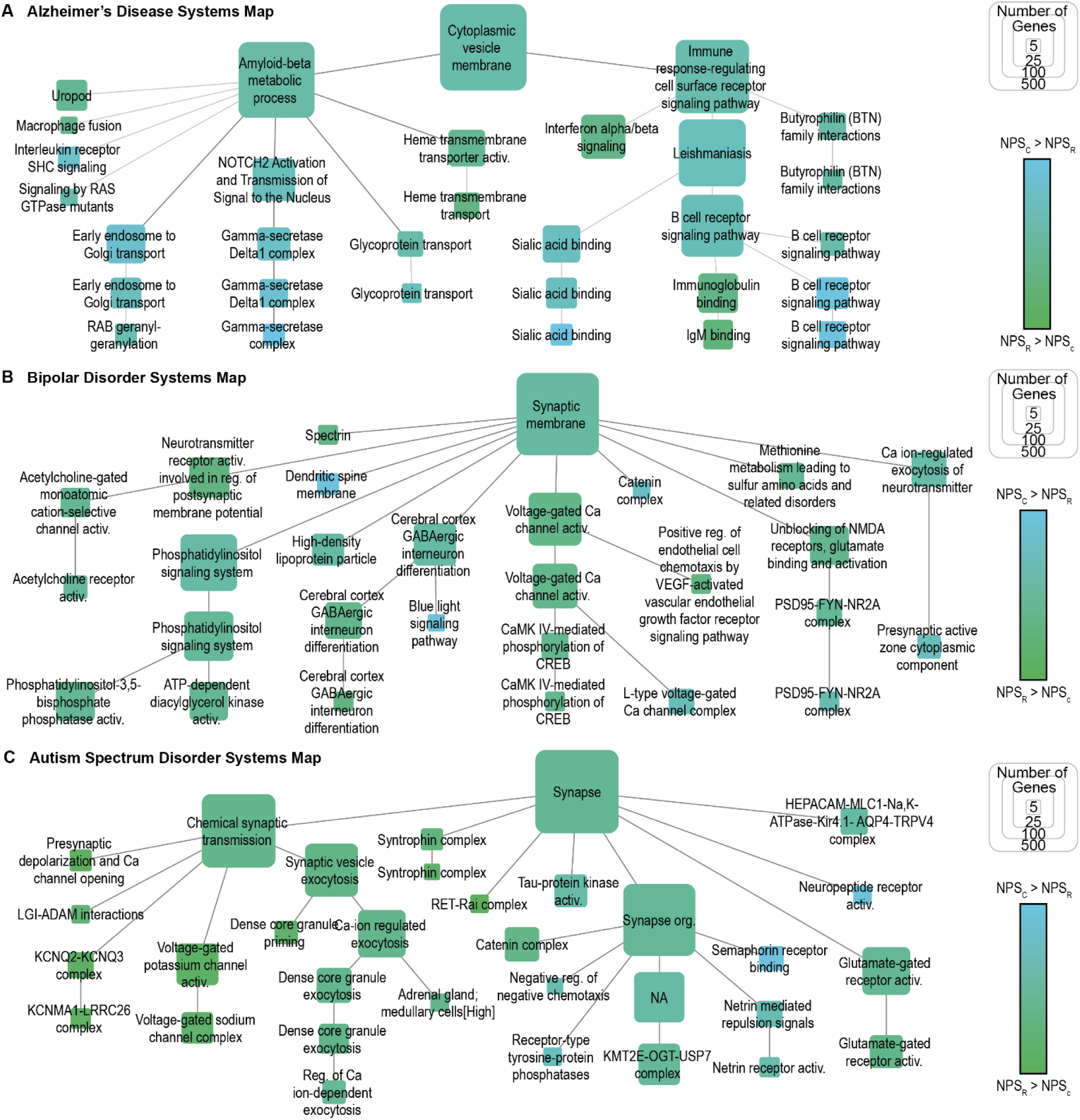
Systems maps for Alzheimer’s disease, bipolar disorder, and autism spectrum disorder. Maps generated via hierarchical community detection of the trait networks and annotated using g:Profiler (Methods). Node size indicates the number of genes per system, and node color represents the ratio of NPSC and NPSR for genes within the system. Systems with more than five genes are displayed. Communities without a significant enrichment were annotated as NA. Related to Figure 6.

**Supplemental Figure 4.**
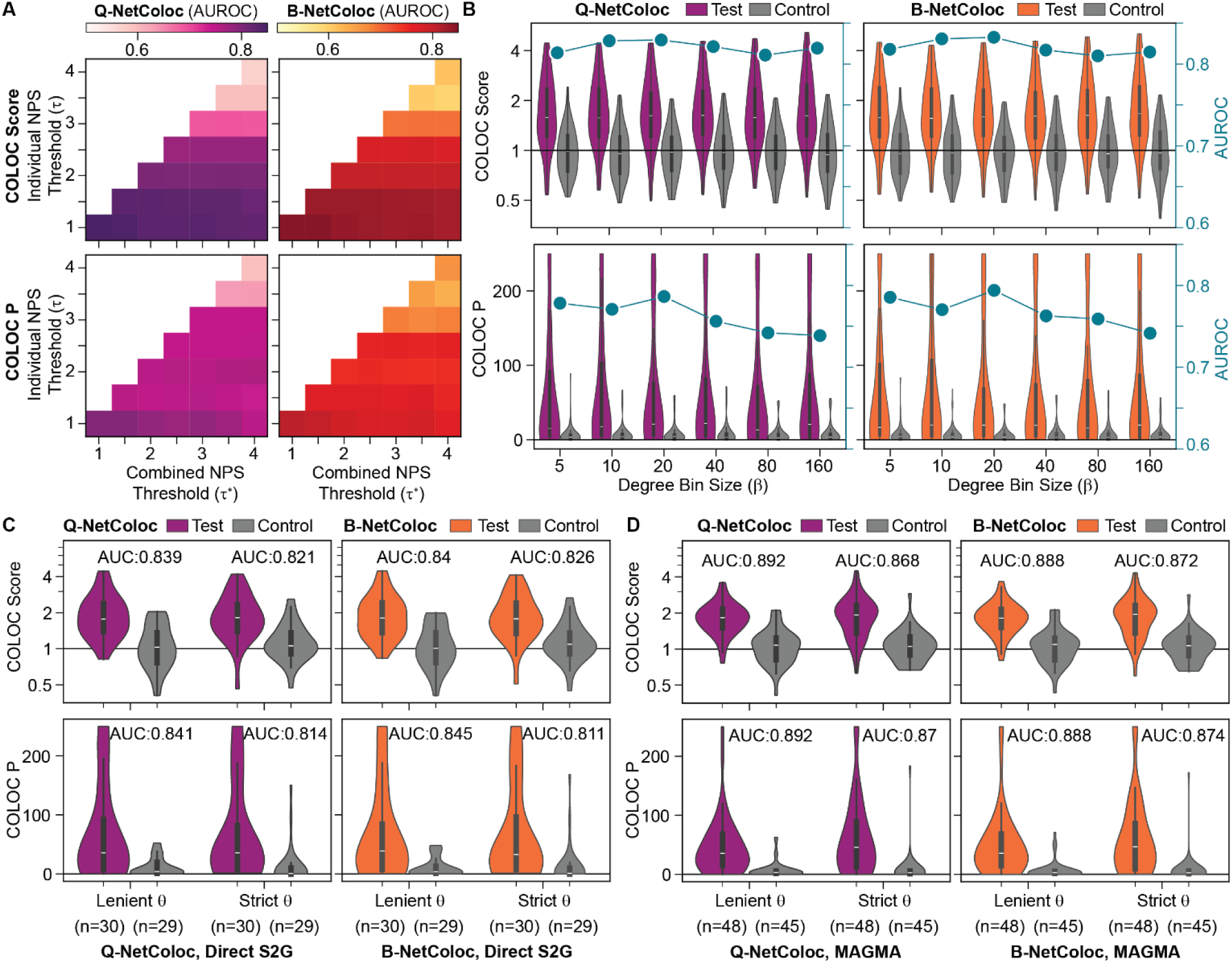
Optimization and benchmarking of network colocalization. Quantitative (Q-NetColoc) and Binary (B-NetColoc) implementations are shown in purple and orange, respectively. All analyses were conducted with partitioned GWAS results for traits not included in the main analysis. Test results are from matched study partitions, while control results are from analysis of mismatched partitions (Methods). **A)** AUROC results for Q-NetColoc and B-NetColoc across different thresholds τ and τ^∗^ for defining the trait network. **B)** Network colocalization results and AUROC values for Q-NetColoc and B-NetColoc across different bin sizes (*β*) used for degree-matched randomization. **C)** Network colocalization results and AUROC values for Q-NetColoc and B-NetColoc at lenient (*p* < 1×10^−5^) and strict (*p* < 1×10^−8^) thresholds for inclusion of trait-associated genes (θ). Trait-associated genes defined by direct SNP-to-Gene (S2G) mapping via the GWAS catalog. **D)** Network colocalization results and AUROC values for Q-NetColoc and B-NetColoc at lenient (*p* < 1×10^−4^) and strict (*p* < 2.5×10^−6^) thresholds for inclusion of trait-associated genes (θ). Trait-associated genes defined by MAGMA gene-level p-values. Related to Methods.

## Notes

### Author Declarations

All human data utilized in this study were openly available before the initiation of the study. Summary results from previously published genome wide association studies (GWAS) were sourced from the GWAS Catalog (https://www.ebi.ac.uk/gwas/api/search/downloads/alternative). Accession numbers for the studies used can be found in Supplemental Table 1. Summary results from previously published gene-level rare variant studies were sourced from the Rare Variant Association Repository (RAVAR, http://www.ravar.bio/api/download/static/gene_fulltable.txt). Publication identifiers (PMIDs) for the studies used can be found in Supplemental Table 1.

